# COVID-19 primary series and booster vaccination and potential for immune imprinting

**DOI:** 10.1101/2022.10.31.22281756

**Authors:** Hiam Chemaitelly, Houssein H. Ayoub, Patrick Tang, Peter V. Coyle, Hadi M. Yassine, Asmaa A. Al Thani, Hebah A. Al-Khatib, Mohammad R. Hasan, Zaina Al-Kanaani, Einas Al-Kuwari, Andrew Jeremijenko, Anvar Hassan Kaleeckal, Ali Nizar Latif, Riyazuddin Mohammad Shaik, Hanan F. Abdul-Rahim, Gheyath K. Nasrallah, Mohamed Ghaith Al-Kuwari, Adeel A. Butt, Hamad Eid Al-Romaihi, Mohamed H. Al-Thani, Abdullatif Al-Khal, Roberto Bertollini, Laith J. Abu-Raddad

## Abstract

Laboratory science evidence suggests possibility of immune imprinting, a negative impact for vaccination on subsequent protective immunity against SARS-CoV-2 infection. We investigated differences in incidence of SARS-CoV-2 reinfection in the cohort of persons who had a primary omicron infection, but different vaccination histories using matched, national, retrospective, cohort studies. Adjusted hazard ratio (AHR) for incidence of reinfection, factoring also adjustment for differences in testing rate, was 0.43 (95% CI: 0.39-0.49) comparing history of two-dose vaccination to no vaccination, 1.47 (95% CI: 1.23-1.76) comparing history of three-dose vaccination to two-dose vaccination, and 0.57 (95% CI: 0.48-0.68) comparing history of three-dose vaccination to no vaccination. Divergence in cumulative incidence curves increased markedly when incidence was dominated by BA.4/BA.5 and BA.2.75* omicron subvariant. History of primary-series vaccination enhanced immune protection against omicron reinfection, but history of booster vaccination compromised protection against omicron reinfection. These findings do not undermine the short-term public health utility of booster vaccination.

**Teaser:** History of booster vaccination showed lower protection against omicron reinfection than history of two-dose vaccination.

## Introduction

Three years into the coronavirus disease 2019 (COVID-19) pandemic, the global population carries heterogenous immune histories derived from various exposures to infection, viral variants, and vaccination (*1*). Laboratory science evidence suggests the possibility of immune imprinting, a negative impact for vaccination on subsequent protective immunity against severe acute respiratory syndrome coronavirus 2 (SARS-CoV-2) induced by vaccination or infection, or a combination of both (*1-4*). Epidemiological evidence for immune imprinting in immune histories related to infection was recently investigated, but no evidence was found for imprinting compromising protection against B.1.1.529 (omicron) subvariants (*5*). A pre-omicron infection followed by an omicron reinfection enhanced protection against a second omicron reinfection (*5*).

We investigated epidemiological evidence for imprinting in immune histories related to vaccination using matched, retrospective cohort studies conducted on the total population of Qatar from onset of the omicron wave on December 19, 2021 (*6*) through September 15, 2022. We compared incidence of SARS-CoV-2 reinfection in the national cohort of individuals who had a primary documented omicron infection after primary-series (two-dose) vaccination (designated as the two-dose cohort) to that in the national cohort of individuals with a documented primary omicron infection, but no vaccination history (designated as the unvaccinated cohort). Analogously, we also compared reinfection incidence in those who had a documented primary omicron infection after booster (third dose) vaccination (designated as the three-dose cohort) to each of the two-dose and unvaccinated cohorts.

These immune histories were investigated because of specific immunological scenarios observed in immunological laboratory data (*1*), because of their pervasiveness in the global population, and because of their potential relevance to the protection of bivalent booster vaccination that is being scaled up in different countries.

A documented primary omicron infection was defined as the first record of a SARS-CoV-2-positive polymerase chain reaction (PCR) or rapid antigen test after onset of the omicron wave in Qatar on December 19, 2021 (*6*) in an individual that had no record of a prior pre-omicron infection. SARS-CoV-2 reinfection was defined, per the conventional definition in the literature, as a documented infection ≥90 days after an earlier infection, to avoid misclassifying prolonged SARS-CoV-2 positivity as reinfection if a shorter time interval is used (*6-8*). Matched pairs were followed from 90 days after the primary omicron infection to record incidence of SARS-CoV-2 reinfection.

## Results

### Two-dose cohort versus unvaccinated cohort

Fig. S1 shows the study population selection process. Table 1 describes baseline characteristics of the full and matched cohorts. Matched cohorts each included 56,802 individuals.

**Table 1.**
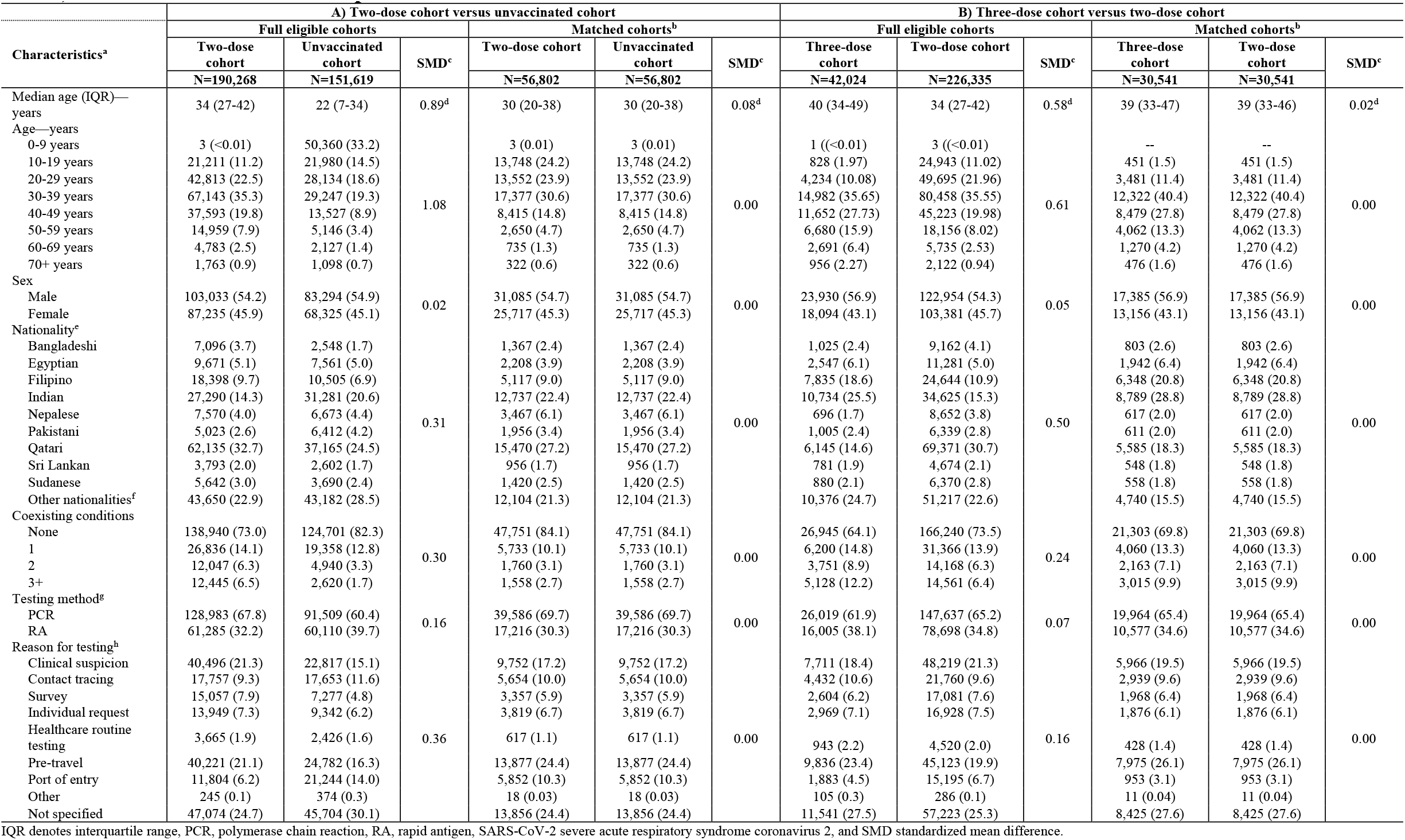
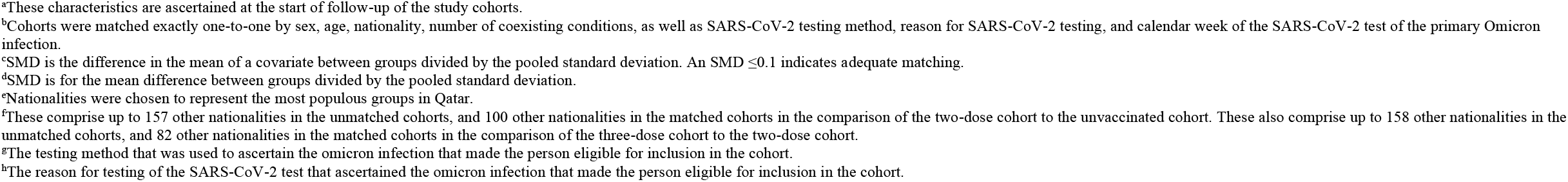
Baseline characteristics of eligible and matched cohorts in studies investigating immune protection against reinfection among those who had a primary infection with an omicron subvariant, but had a history of A) two-dose vaccination compared to no vaccination, and B) three-dose vaccination compared to two-dose vaccination.

| Characteristics<sup>a</sup> | A) Two-dose cohort versus unvaccinated cohort |  |  |  |  |  | B) Three-dose cohort versus two-dose cohort |  |  |  |  |  |
| --- | --- | --- | --- | --- | --- | --- | --- | --- | --- | --- | --- | --- |
|  | Full eligible cohorts |  |  | Matched cohorts<sup>b</sup> |  |  | Full eligible cohorts |  |  | Matched cohorts<sup>b</sup> |  |  |
|  | Two-dose cohort | Unvaccinated cohort | SMD<sup>c</sup> | Two-dose cohort | Unvaccinated cohort | SMD<sup>c</sup> | Three-dose cohort | Two-dose cohort | SMD<sup>c</sup> | Three-dose cohort | Two-dose cohort | SMD<sup>c</sup> |
|  | N=190,268 | N=151,619 |  | N=56,802 | N=56,802 |  | N=42,024 | N=226,335 |  | N=30,541 | N=30,541 |  |
| Median age (IQR)—years | 34 (27-42) | 22 (7-34) | 0.89<sup>d</sup> | 30 (20-38) | 30 (20-38) | 0.08<sup>d</sup> | 40 (34-49) | 34 (27-42) | 0.58<sup>d</sup> | 39 (33-47) | 39 (33-46) | 0.02<sup>d</sup> |
| Age—years |  |  |  |  |  |  |  |  |  |  |  |  |
| 0-9 years | 3 (<0.01) | 50,360 (33.2) |  | 3 (0.01) | 3 (0.01) |  | 1 (<0.01) | 3 (<0.01) |  | -- | -- |  |
| 10-19 years | 21,211 (11.2) | 21,980 (14.5) |  | 13,748 (24.2) | 13,748 (24.2) |  | 828 (1.97) | 24,943 (11.02) |  | 451 (1.5) | 451 (1.5) |  |
| 20-29 years | 42,813 (22.5) | 28,134 (18.6) |  | 13,552 (23.9) | 13,552 (23.9) |  | 4,234 (10.08) | 49,695 (21.96) |  | 3,481 (11.4) | 3,481 (11.4) |  |
| 30-39 years | 67,143 (35.3) | 29,247 (19.3) |  | 17,377 (30.6) | 17,377 (30.6) |  | 14,982 (35.65) | 80,458 (35.55) |  | 12,322 (40.4) | 12,322 (40.4) |  |
| 40-49 years | 37,593 (19.8) | 13,527 (8.9) | 1.08 | 8,415 (14.8) | 8,415 (14.8) | 0.00 | 11,652 (27.73) | 45,223 (19.98) | 0.61 | 8,479 (27.8) | 8,479 (27.8) | 0.00 |
| 50-59 years | 14,959 (7.9) | 5,146 (3.4) |  | 2,650 (4.7) | 2,650 (4.7) |  | 6,680 (15.9) | 18,156 (8.02) |  | 4,062 (13.3) | 4,062 (13.3) |  |
| 60-69 years | 4,783 (2.5) | 2,127 (1.4) |  | 735 (1.3) | 735 (1.3) |  | 2,691 (6.4) | 5,735 (2.53) |  | 1,270 (4.2) | 1,270 (4.2) |  |
| 70+ years | 1,763 (0.9) | 1,098 (0.7) |  | 322 (0.6) | 322 (0.6) |  | 956 (2.27) | 2,122 (0.94) |  | 476 (1.6) | 476 (1.6) |  |
| Sex |  |  |  |  |  |  |  |  |  |  |  |  |
| Male | 103,033 (54.2) | 83,294 (54.9) |  | 31,085 (54.7) | 31,085 (54.7) |  | 23,930 (56.9) | 122,954 (54.3) |  | 17,385 (56.9) | 17,385 (56.9) |  |
| Female | 87,235 (45.9) | 68,325 (45.1) | 0.02 | 25,717 (45.3) | 25,717 (45.3) | 0.00 | 18,094 (43.1) | 103,381 (45.7) | 0.05 | 13,156 (43.1) | 13,156 (43.1) | 0.00 |
| Nationality<sup>e</sup> |  |  |  |  |  |  |  |  |  |  |  |  |
| Bangladeshi | 7,096 (3.7) | 2,548 (1.7) |  | 1,367 (2.4) | 1,367 (2.4) |  | 1,025 (2.4) | 9,162 (4.1) |  | 803 (2.6) | 803 (2.6) |  |
| Egyptian | 9,671 (5.1) | 7,561 (5.0) |  | 2,208 (3.9) | 2,208 (3.9) |  | 2,547 (6.1) | 11,281 (5.0) |  | 1,942 (6.4) | 1,942 (6.4) |  |
| Filipino | 18,398 (9.7) | 10,505 (6.9) |  | 5,117 (9.0) | 5,117 (9.0) |  | 7,835 (18.6) | 24,644 (10.9) |  | 6,348 (20.8) | 6,348 (20.8) |  |
| Indian | 27,290 (14.3) | 31,281 (20.6) |  | 12,737 (22.4) | 12,737 (22.4) |  | 10,734 (25.5) | 34,625 (15.3) |  | 8,789 (28.8) | 8,789 (28.8) |  |
| Nepalese | 7,570 (4.0) | 6,673 (4.4) |  | 3,467 (6.1) | 3,467 (6.1) |  | 696 (1.7) | 8,652 (3.8) |  | 617 (2.0) | 617 (2.0) |  |
| Pakistani | 5,023 (2.6) | 6,412 (4.2) | 0.31 | 1,956 (3.4) | 1,956 (3.4) | 0.00 | 1,005 (2.4) | 6,339 (2.8) | 0.50 | 611 (2.0) | 611 (2.0) | 0.00 |
| Qatari | 62,135 (32.7) | 37,165 (24.5) |  | 15,470 (27.2) | 15,470 (27.2) |  | 6,145 (14.6) | 69,371 (30.7) |  | 5,585 (18.3) | 5,585 (18.3) |  |
| Sri Lankan | 3,793 (2.0) | 2,602 (1.7) |  | 956 (1.7) | 956 (1.7) |  | 781 (1.9) | 4,674 (2.1) |  | 548 (1.8) | 548 (1.8) |  |
| Sudanese | 5,642 (3.0) | 3,690 (2.4) |  | 1,420 (2.5) | 1,420 (2.5) |  | 880 (2.1) | 6,370 (2.8) |  | 558 (1.8) | 558 (1.8) |  |
| Other nationalities<sup>f</sup> | 43,650 (22.9) | 43,182 (28.5) |  | 12,104 (21.3) | 12,104 (21.3) |  | 10,376 (24.7) | 51,217 (22.6) |  | 4,740 (15.5) | 4,740 (15.5) |  |
| Coexisting conditions |  |  |  |  |  |  |  |  |  |  |  |  |
| None | 138,940 (73.0) | 124,701 (82.3) |  | 47,751 (84.1) | 47,751 (84.1) |  | 26,945 (64.1) | 166,240 (73.5) |  | 21,303 (69.8) | 21,303 (69.8) |  |
| 1 | 26,836 (14.1) | 19,358 (12.8) |  | 5,733 (10.1) | 5,733 (10.1) |  | 6,200 (14.8) | 31,366 (13.9) |  | 4,060 (13.3) | 4,060 (13.3) |  |
| 2 | 12,047 (6.3) | 4,940 (3.3) | 0.30 | 1,760 (3.1) | 1,760 (3.1) | 0.00 | 3,751 (8.9) | 14,168 (6.3) | 0.24 | 2,163 (7.1) | 2,163 (7.1) | 0.00 |
| 3+ | 12,445 (6.5) | 2,620 (1.7) |  | 1,558 (2.7) | 1,558 (2.7) |  | 5,128 (12.2) | 14,561 (6.4) |  | 3,015 (9.9) | 3,015 (9.9) |  |
| Testing method<sup>g</sup> |  |  |  |  |  |  |  |  |  |  |  |  |
| PCR | 128,983 (67.8) | 91,509 (60.4) |  | 39,586 (69.7) | 39,586 (69.7) |  | 26,019 (61.9) | 147,637 (65.2) |  | 19,964 (65.4) | 19,964 (65.4) |  |
| RA | 61,285 (32.2) | 60,110 (39.7) | 0.16 | 17,216 (30.3) | 17,216 (30.3) | 0.00 | 16,005 (38.1) | 78,698 (34.8) | 0.07 | 10,577 (34.6) | 10,577 (34.6) | 0.00 |
| Reason for testing<sup>h</sup> |  |  |  |  |  |  |  |  |  |  |  |  |
| Clinical suspicion | 40,496 (21.3) | 22,817 (15.1) |  | 9,752 (17.2) | 9,752 (17.2) |  | 7,711 (18.4) | 48,219 (21.3) |  | 5,966 (19.5) | 5,966 (19.5) |  |
| Contact tracing | 17,757 (9.3) | 17,653 (11.6) |  | 5,654 (10.0) | 5,654 (10.0) |  | 4,432 (10.6) | 21,760 (9.6) |  | 2,939 (9.6) | 2,939 (9.6) |  |
| Survey | 15,057 (7.9) | 7,277 (4.8) |  | 3,357 (5.9) | 3,357 (5.9) |  | 2,604 (6.2) | 17,081 (7.6) |  | 1,968 (6.4) | 1,968 (6.4) |  |
| Individual request | 13,949 (7.3) | 9,342 (6.2) |  | 3,819 (6.7) | 3,819 (6.7) |  | 2,969 (7.1) | 16,928 (7.5) |  | 1,876 (6.1) | 1,876 (6.1) |  |
| Healthcare routine testing | 3,665 (1.9) | 2,426 (1.6) | 0.36 | 617 (1.1) | 617 (1.1) | 0.00 | 943 (2.2) | 4,520 (2.0) | 0.16 | 428 (1.4) | 428 (1.4) | 0.00 |
| Pre-travel | 40,221 (21.1) | 24,782 (16.3) |  | 13,877 (24.4) | 13,877 (24.4) |  | 9,836 (23.4) | 45,123 (19.9) |  | 7,975 (26.1) | 7,975 (26.1) |  |
| Port of entry | 11,804 (6.2) | 21,244 (14.0) |  | 5,852 (10.3) | 5,852 (10.3) |  | 1,883 (4.5) | 15,195 (6.7) |  | 953 (3.1) | 953 (3.1) |  |
| Other | 245 (0.1) | 374 (0.3) |  | 18 (0.03) | 18 (0.03) |  | 105 (0.3) | 286 (0.1) |  | 11 (0.04) | 11 (0.04) |  |
| Not specified | 47,074 (24.7) | 45,704 (30.1) |  | 13,856 (24.4) | 13,856 (24.4) |  | 11,541 (27.5) | 57,223 (25.3) |  | 8,425 (27.6) | 8,425 (27.6) |  |

IQR denotes interquartile range, PCR, polymerase chain reaction, RA, rapid antigen, SARS-CoV-2 severe acute respiratory syndrome coronavirus 2, and SMD standardized mean difference.
<sup>a</sup>These characteristics are ascertained at the start of follow-up of the study cohorts.
<sup>b</sup>Cohorts were matched exactly one-to-one by sex, age, nationality, number of coexisting conditions, as well as SARS-CoV-2 testing method, reason for SARS-CoV-2 testing, and calendar week of the SARS-CoV-2 test of the primary Omicron infection.
<sup>c</sup>SMD is the difference in the mean of a covariate between groups divided by the pooled standard deviation. An $SMD \leq 0.1$ indicates adequate matching.
<sup>d</sup>SMD is for the mean difference between groups divided by the pooled standard deviation.
<sup>e</sup>Nationalities were chosen to represent the most populous groups in Qatar.
<sup>f</sup>These comprise up to 157 other nationalities in the unmatched cohorts, and 100 other nationalities in the matched cohorts in the comparison of the two-dose cohort to the unvaccinated cohort. These also comprise up to 158 other nationalities in the unmatched cohorts, and 82 other nationalities in the matched cohorts in the comparison of the three-dose cohort to the two-dose cohort.
<sup>g</sup>The testing method that was used to ascertain the omicron infection that made the person eligible for inclusion in the cohort.
<sup>h</sup>The reason for testing of the SARS-CoV-2 test that ascertained the omicron infection that made the person eligible for inclusion in the cohort.

Median date of the second vaccine dose for the two-dose cohort was June 9, 2021. Median duration between the second dose and start of follow-up was 312 days (interquartile range (IQR), 264-352 days). Median duration of follow-up was 157 days (IQR, 140-164 days) for the two-dose cohort and 157 days (IQR, 139-164 days) for the unvaccinated cohort (Fig. 1A). There were 573 reinfections in the two-dose cohort and 1,044 reinfections in the unvaccinated cohort during follow-up (Fig. S1). None progressed to severe, critical, or fatal COVID-19.

**Fig. 1.**
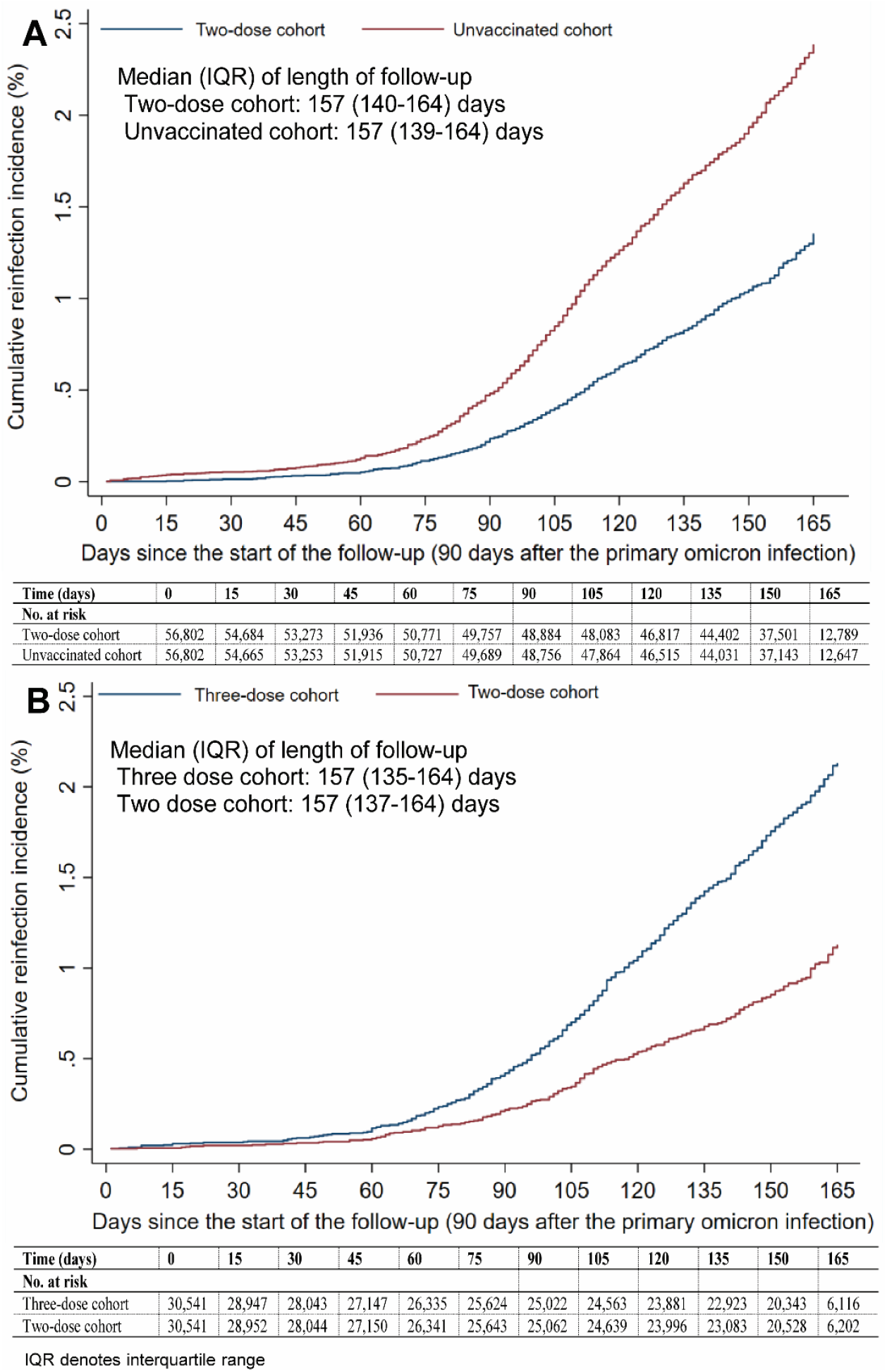
Cumulative incidence of reinfection among those who had a primary infection with an omicron subvariant after A) two-dose vaccination compared to no vaccination, and B) three-dose vaccination compared to two-dose vaccination using the Kaplan-Meier estimator.

Cumulative incidence of reinfection was 1.4% (95% CI: 1.2-1.5%) for the two-dose cohort and 2.4% (95% CI: 2.2-2.5%) for the unvaccinated cohort, after 165 days of follow-up (Fig. 1A). In the first 70 days of follow-up, incidence was dominated by BA.2 (*9-11*). Subsequently, incidence was dominated by BA.4/BA.5 (*12*), and then by BA.2.75* (*13*) (predominantly BA.2.75.2). Divergence between the cumulative incidence curves increased markedly when incidence was no longer dominated by BA.2.

The hazard ratio comparing incidence of reinfection in the two-dose cohort to that in the unvaccinated cohort, adjusted for matching factors, was 0.59 (95% CI: 0.53-0.67; Table 2). The adjusted hazard ratio appeared stable by month of follow-up (Fig. 2A). The proportion of individuals who had a test during follow-up was 48.9% for the two-dose cohort and 37.0% for the unvaccinated cohort. The testing frequency was 0.93 and 0.67 tests per person, respectively. Adjusting the hazard ratio additionally for differences in testing rate between cohorts yielded an adjusted hazard ratio of 0.43 (95% CI: 0.39-0.49).

**Table 2.** Hazard ratios for incidence of SARS-CoV-2 reinfection in studies investigating immune protection among those who had a primary infection with an omicron subvariant, but different vaccination histories.

| Epidemiological measure | Cohorts<sup>a</sup> |  |
| --- | --- | --- |
|  | Two-dose cohort | Unvaccinated cohort |
| **Two-dose vaccination versus no vaccination before primary omicron infection** |  |  |
| Incident reinfections (n) | 573 | 1,044 |
| Total follow-up time (person-weeks) | 1,124,759 | 1,121,092 |
| Incidence rate of reinfection (per 10,000 person-weeks; 95% CI) | 5.1 (4.7 to 5.5) | 9.3 (8.8 to 9.9) |
| Unadjusted hazard ratio for SARS-CoV-2 reinfection (95% CI) | 0.55 (0.49 to 0.60) |  |
| Adjusted hazard ratio for SARS-CoV-2 reinfection (95% CI)<sup>b</sup> | 0.59 (0.53 to 0.67) |  |
| Hazard ratio for SARS-CoV-2 reinfection additionally adjusted for differences in testing rate (95% CI)<sup>b</sup> | 0.43 (0.39 to 0.49) |  |
| **Three-dose vaccination versus two-dose vaccination before primary omicron infection** | **Three-dose cohort** | **Two-dose cohort** |
| Incident reinfections (n) | 480 | 248 |
| Total follow-up time (person-weeks) | 585,068 | 586,527 |
| Incidence rate of reinfection (per 10,000 person-weeks; 95% CI) | 8.2 (7.5 to 9.0) | 4.2 (3.7 to 4.8) |
| Unadjusted hazard ratio for SARS-CoV-2 reinfection (95% CI) | 1.94 (1.67 to 2.27) |  |
| Adjusted hazard ratio for SARS-CoV-2 reinfection (95% CI)<sup>b</sup> | 1.96 (1.64 to 2.34) |  |
| Hazard ratio for SARS-CoV-2 reinfection additionally adjusted for differences in testing rate (95% CI)<sup>b</sup> | 1.47 (1.23 to 1.76) |  |
| **Three-dose vaccination versus no vaccination before primary omicron infection** | **Three-dose cohort** | **Unvaccinated cohort** |
| Incident reinfections (n) | 337 | 323 |
| Total follow-up time (person-weeks) | 397,179 | 396,929 |
| Incidence rate of reinfection (per 10,000 person-weeks; 95% CI) | 8.5 (7.6 to 9.4) | 8.1 (7.3 to 9.1) |
| Unadjusted hazard ratio for SARS-CoV-2 reinfection (95% CI) | 1.04 (0.89 to 1.21) |  |
| Adjusted hazard ratio for SARS-CoV-2 reinfection (95% CI)<sup>b</sup> | 1.10 (0.92 to 1.31) |  |
| Hazard ratio for SARS-CoV-2 reinfection additionally adjusted for differences in testing rate (95% CI)<sup>b</sup> | 0.57 (0.48 to 0.68) |  |

CI denotes confidence interval and SARS-CoV-2 severe acute respiratory syndrome coronavirus 2.
<sup>a</sup>Cohorts were matched exactly one-to-one by sex, age, nationality, number of coexisting conditions, as well as SARS-CoV-2 testing method, reason for SARS-CoV-2 testing, and calendar week of the SARS-CoV-2 test of the primary omicron infection.
<sup>b</sup>Cox regression analysis adjusted for sex, 10-year age groups, 10 nationality groups, number of coexisting conditions, as well as SARS-CoV-2 testing method, reason for SARS-CoV-2 testing, and calendar week of the SARS-CoV-2 test of the primary omicron infection.

**Fig. 2.**
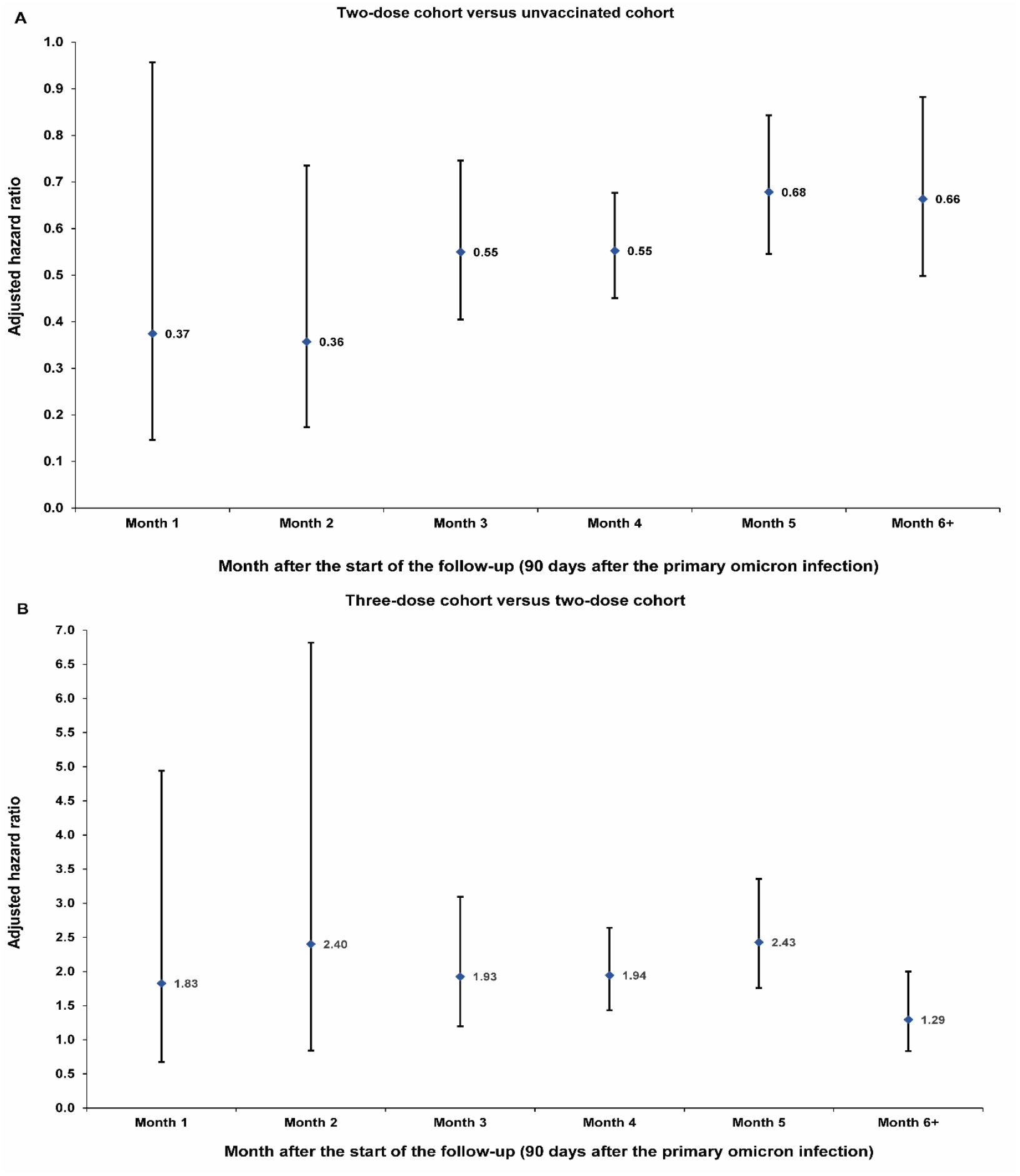
Adjusted hazard ratio by month of follow-up for SARS-CoV-2 reinfection among those who had a primary infection with an omicron subvariant A) after two-dose vaccination compared to no vaccination, and B) after three-dose vaccination compared to two-dose vaccination. Analyses were performed on 56,802 and 30,541 matched pairs, respectively. Error bars indicate 95% confidence intervals.

### Three-dose cohort versus two-dose cohort

Fig. S2 shows the study population selection process. Table 1 describes baseline characteristics of the full and matched cohorts. Matched cohorts each included 30,541 individuals.

Median dates of the second and third vaccine doses for the three-dose cohort were March 26, 2021 and December 6, 2021, respectively. Median date of the second vaccine dose for the two-dose cohort was May 11, 2021. Median duration between the third dose and start of follow-up was 124 days (IQR, 103-143 days), and between the second dose and start of follow-up was 334 days (IQR, 286-371 days). Median duration of follow-up was 157 days (IQR, 135-164 days) in the three-dose cohort and 157 days (IQR, 137-164 days) in the two-dose cohort (Fig. 1B). There were 480 reinfections in the three-dose cohort and 248 reinfections in the two-dose cohort during follow-up (Fig. S2). None progressed to severe, critical, or fatal COVID-19.

Cumulative incidence of reinfection was 2.1% (95% CI: 1.9-2.3%) for the three-dose cohort and 1.1% (95% CI: 1.0-1.3%) for the two-dose cohort, after 165 days of follow-up (Fig. 1B). In the first 70 days of follow-up, incidence was dominated by BA.2 (*9-11*). Subsequently, incidence was dominated by BA.4/BA.5 (*12*), and then by BA.2.75* (*13*). Divergence between the cumulative incidence curves increased markedly when incidence was no longer dominated by BA.2.

The adjusted hazard ratio comparing incidence of reinfection in the three-dose cohort to that in the two-dose cohort was 1.96 (95% CI: 1.64-2.34; Table 2). The adjusted hazard ratio appeared stable by month of follow-up, but with wide 95% confidence intervals (Fig. 2B). The proportion of individuals who had a test during follow-up was 63.1% for the three-dose cohort and 49.0% for the two-dose cohort. The testing frequency was 1.39 and 0.98 tests per person, respectively. Adjusting the hazard ratio additionally for differences in testing rate between cohorts yielded an adjusted hazard ratio of 1.47 (95% CI: 1.23-1.76).

In the first sensitivity analysis with the cohorts being matched by the Charlson comorbidity index, instead of the number of chronic coexisting conditions, the adjusted hazard ratio, including also the adjustment for the differences in testing rate, was 1.39 (95% CI: 1.16-1.67) (Table S1).

In the second sensitivity analysis with the cohorts being matched additionally by primary-series vaccine type (two doses of BNT162b2 or two doses of mRNA-1273), the adjusted hazard ratio, including also the adjustment for the differences in testing rate, was 1.43 (95% CI: 1.19-1.71) (Table S1). In the subgroup analysis including only BNT162b2-vaccinated individuals, the adjusted hazard ratio was 1.39 (95% CI: 1.15-1.68). In the subgroup analysis including only mRNA-1273-vaccinated individuals, the adjusted hazard ratio was 1.83 (95% CI: 1.03-3.28).

### Three-dose cohort versus unvaccinated cohort

Fig. S3 shows the study population selection process. Table S2 describes baseline characteristics of the full and matched cohorts. Cumulative incidence of reinfection is shown in Fig. S4A.

The adjusted hazard ratio comparing incidence of reinfection in the three-dose cohort to that in the unvaccinated cohort was 1.10 (95% CI: 0.92-1.31; Table 2). The adjusted hazard ratio appeared stable by month of follow-up, but with wide 95% confidence intervals (Fig. S4B). The proportion of individuals who had a test during follow-up was 66.4% for the three-dose cohort and 36.8% for the unvaccinated cohort. The testing frequency was 1.46 and 0.70 tests per person, respectively. Adjusting the hazard ratio additionally for differences in testing rate between cohorts yielded an adjusted hazard ratio of 0.57 (95% CI: 0.48-0.68).

The results of this additional study confirm the relative differences in incidence of reinfection observed in the first two studies, with incidence being lowest among the two-dose cohort and highest among the unvaccinated cohort.

## Discussion

Primary-series vaccination followed by a primary omicron infection was associated with enhanced immune protection against omicron reinfection compared to primary omicron infection with no prior vaccination. This result is striking because the start of follow-up in this study was ∼1 year after the two-dose primary series. Protection of the primary series against omicron infection that is mediated by neutralizing antibodies should have fully waned by this time, considering how rapidly vaccine protection wanes against omicron subvariants (*10, 14*). This finding suggests that the primary omicron infection may have stimulated other components of the immune system, specifically immune memory of the earlier primary-series immune response in a manner that enhanced protection against a subsequent omicron reinfection, particularly against BA.4/BA.5 and BA.2.75*.

Remarkably, similar effect and effect size were observed recently in an analogous study (*5*). Incidence of reinfection among unvaccinated persons who had contracted an omicron infection following an earlier pre-omicron infection was lower than incidence of reinfection among unvaccinated persons who had only an omicron infection and no prior pre-omicron infection (*5*). mRNA vaccines used in Qatar are based on index-virus design (*15, 16*). The median duration between the first and second vaccine doses was <1 month (*17*). Given this short duration between doses, two-dose vaccination counts perhaps as a single pre-omicron immunological event. This may explain the similarity in both effect and effect size in these two studies, since in essence, both investigate immune protection elicited by a pre-omicron immunological event followed by an omicron immunological event, compared to protection of only a single omicron event.

While two-dose vaccination was associated with enhanced protection against subsequent omicron reinfection, three-dose vaccination was associated with reduced protection compared to that of two-dose vaccination. This finding suggests that the immune response against the primary omicron infection may have been compromised by differential immune imprinting in those who received a third booster dose, apparently consistent with laboratory science data (*1-4*) and emerging epidemiologic data (*18-21*) on imprinting effects. The booster dose, a pre-omicron immunological event, that occurred several months after the primary-series vaccination, another pre-omicron immunological event, may have trained the immune response to expect a specific narrow pre-omicron challenge; thus, the response was inferior when the actual challenge was an immune-evasive omicron subvariant. Repeat immunological events of the same kind (here pre-omicron challenge) may be associated with compromised protection against a new kind of immunological event (here omicron challenge).

This imprinting effect appears related to the memory component of the immune response, perhaps explaining why the effect was observed only after waning of the antibody-mediated short-term booster protection, as supported also by another study on the same population of the long-term effectiveness of booster vaccination (*20*). Those with a booster may have had their immune memory geared and narrowed down toward expecting a specific pre-omicron challenge (*22*). The imprinting effect seems to arise from the mismatch between such specific immune memory and the actual substantially different immune challenge (*22*). The size of the imprinting effect appeared also to be larger for mRNA-1273-vaccinated persons than for BNT162b2-vaccinated persons, possibly because of the larger dose of the mRNA-1273 vaccine (*17*), and perhaps suggesting a dose-response relationship for the imprinting effect.

We investigated two immune histories with different effects for immune imprinting on each. Primary-series vaccination followed by a primary omicron infection enhanced immune protection against omicron reinfection. Booster vaccination followed by a primary omicron infection compromised protection against omicron reinfection. This highlights the complexity of the immunity landscape at this stage of the pandemic, in which people have different immune histories. These findings, however, do not undermine the utility of booster vaccination, at least in the short-term. Compromised protection was observed only after waning of the antibody-mediated short-term booster protection, as follow-up commenced >4 months after the booster, at a time when booster effectiveness is expected to be marginal (*10, 14, 20*). There is no question that the booster dose reduced infection incidence in the first 6 months after its administration, based on evidence from this same population (*9, 10, 20, 23*). Nonetheless, the findings suggest that short-term effects of boosters may differ from their long-term effects.

Although we planned to investigate effectiveness against severe COVID-19, no reinfection in any cohort of the three studies progressed to severe, critical, or fatal COVID-19. Though some of the COVID-19 patients were hospitalized, none reached the World Health Organization classification of severe or critical COVID-19, and none ended up with COVID-19 death following the longitudinal review of their individual charts. This outcome is not unexpected given the lower severity of omicron infections (*24, 25*) and the strong protection of natural infection against severe COVID-19 at reinfection, estimated at 97% in this same population (*26*), as well as the long-term effectiveness of primary-series and boosters against severe COVID-19 (*9, 10, 14, 20, 27, 28*). While we were unable to quantify effects of immune imprinting on COVID-19 severity, the results do not suggest imprinting compromising protection against severe COVID-19. This has also been supported by another analysis on the same population (*20*).

The central analysis in this study compares incidence of infection among boosted persons versus those with only a primary series, both groups of which had an omicron primary infection after vaccination. However, these two groups may not be immunologically comparable with respect to their ability to produce a strong immune response following vaccination and omicron infection. The three-dose group consists of individuals with three vaccine doses and a primary infection shortly after the third dose. By contrast, the two-dose group consists of individuals with only two vaccine doses and a primary infection long after their second dose. It is possible that the shorter duration between dose and omicron infection in the three-dose group versus the two-dose group may have contributed to inferior immunological response to the omicron infection, perhaps explaining the higher incidence among boosted persons thereafter. However, the negative imprinting effect observed in this study has now been also observed among groups who are immunologically comparable with respect to their ability to produce a strong immune response following vaccination and/or infection (*20*) arguing against this explanation of the study results.

Following the preprint of this article (*29*) it has been suggested that the conditioning on having infection may introduce bias that explains the higher incidence among boosted persons (*30*). Since the groups have different immune histories prior to primary infection, with one history more protective than the other one, the conditioning on having the infection may implicitly select for persons with more propensity for infection in the group that had the more protective immune history prior to the primary infection. Persons in the three-dose group may have chosen to receive a third vaccine dose because they are aware that they have high levels of exposure, thereby also explaining the higher infection incidence among boosted persons.

However, if this bias existed, its effect needs to be consistent throughout the time of follow-up, not only in one part of it as opposed to another. The results of the analyses presented here, and the earlier analysis for natural immunity (*5*), are not consistent with such a bias effect. There were no differences in incidence between the groups when incidence was due to BA.1/BA.2. The differences between the groups were observed only after incidence was dominated by BA.4/BA.5, consistent with an immune imprinting effect rather than a bias effect.

Moreover, in the analysis comparing history of pre-omicron infection to no pre-omicron infection (*5*), and in the analysis comparing history of primary-series vaccination to no vaccination, a strong positive imprinting effect was found, opposite in direction to effect of this potential bias. If bias existed, the already strong positive imprinting effect is substantially underestimated, an outcome that does not seem plausible given how strong the effect was already in the opposite direction. The effect size was also similar for both of these analyses, despite the differences in immune history, further supporting immune imprinting as an explanation of the study outcomes. Lastly, rigorous matching was implemented to balance infection exposure risk across the groups, and this may have minimized the effect of bias.

This study has limitations. We investigated incidence of documented reinfections, but undocumented reinfections may have occurred. Unvaccinated individuals are a minority in Qatar, and may not be truly immune-naïve due to undocumented prior infections or undocumented vaccinations, perhaps outside the country, especially now that we are three years into this pandemic. Bias due to unequal depletion of the unvaccinated versus vaccinated susceptible population may underestimate vaccine protection (*31*). With Qatar’s young population, our findings may not be generalizable to older individuals or to other countries where elderly citizens constitute a large proportion of the total population.

Testing rate differed between cohorts suggesting the possibility of bias due to differential outcome ascertainment. Receiving a booster dose could be correlated with health-seeking behavior that would result in more frequent testing. Different travel testing guidelines for vaccinated and unvaccinated individuals affect also the testing rate. Such bias due to testing differences may affect the estimated effects and may explain the higher infection incidence among boosted persons. However, the adjustment for the differences in testing rate showed overall similar findings to the main-analysis findings. While the adjustment quantitatively affected the estimated hazard ratios, the adjusted analyses confirmed the finding of higher incidence among boosted persons compared to those with only a primary series. Of note also that the study matched observable confounders across cohorts to control for potential effects of differentials in testing across confounder values. The ratio of testing frequency in the matched cohorts was also overall stable over time of follow-up suggesting absence of substantial differential changes in behavior over time (Fig. S5). Therefore, bias due to differences in testing may not explain the negative imprinting effect observed in this study.

Home-based rapid antigen testing is not documented in Qatar, and is not factored in these analyses. However, there is no reason to believe that home-based testing could have differentially affected the followed cohorts to alter study estimates. Matching was done while factoring key socio-demographic characteristics of the population (*32-36*), such as nationality, age, and sex, and this may also have controlled or reduced differences in home-based testing between cohorts. Nationality, age, and sex provide a powerful proxy for socio-economic status in Qatar (*32-36*). Nationality is also strongly associated with occupation (*32, 34-36*).

Comorbidities were ascertained and classified based on the ICD-10 codes for chronic conditions as recorded in the electronic health record encounters of each individual in the Cerner-system national database that includes all citizens and residents registered in the national and universal public healthcare system. Individuals who have comorbidities but never sought care in the public healthcare system, or seek care exclusively in private healthcare facilities, were classified as individuals with no comorbidity due to absence of recorded encounters for them. This misclassification bias is not likely to have affected the study results considering that the proportion of persons with serious coexisting conditions is small in the predominantly young and working-age population of Qatar (*32, 37*). The national list of vaccine prioritization included only 19,800 individuals of all age groups with serious co-morbid conditions to be prioritized in the first phase of vaccine roll-out (*27*). Of note that the results were invariable by matching by the Charlson comorbidity index instead of the number of coexisting conditions.

As an observational study, investigated cohorts were neither blinded nor randomized, so unmeasured or uncontrolled confounding cannot be excluded. Although matching covered key factors affecting infection exposure (*32-36*), it was not possible for other factors such as geography or occupation, for which data were unavailable. However, Qatar is essentially a city state and infection incidence was broadly distributed across neighborhoods. Nearly 90% of Qatar’s population are expatriates from over 150 countries, who come here for employment (*32*). Nationality, age, and sex provide a powerful proxy for socio-economic status and occupation in this country (*32-36*).

The matching prescription used in this study was investigated in previous studies of different epidemiologic designs, and using control groups to test for null effects (*17, 27, 28, 38, 39*). These control groups included unvaccinated cohorts versus vaccinated cohorts within two weeks of the first dose (*27, 28, 38, 39*), when vaccine protection is negligible (*15, 16*), and mRNA-1273-versus BNT162b2-vaccinated cohorts, also in the first two weeks after the first dose (*17*). These studies showed repeatedly and at different times during the pandemic that this prescription provides adequate control of differences in infection exposure (*17, 27, 28, 38, 39*), suggesting that the employed matching may also have controlled for differences in infection exposure in the present analyses. All analyses were implemented on Qatar’s total population, perhaps minimizing the likelihood of bias.

In conclusion, primary-series vaccination followed by a primary omicron infection enhanced immune protection against omicron reinfection. However, booster vaccination followed by a primary omicron infection compromised protection against omicron reinfection, perhaps because it involved repeat pre-omicron immunological events that are mismatched with currently circulating SARS-CoV-2 variants. These findings do not undermine the utility of booster vaccination in the short-term, but may point to potential complexities in designing boosters with optimal effects.

## Materials and Methods

### Study population and data sources

This study was conducted in the population of Qatar from onset of the omicron wave on December 19, 2021 (*6*) through September 15, 2022. It analyzed the national, federated databases for coronavirus disease 2019 (COVID-19) laboratory testing, vaccination, hospitalization, and death, retrieved from the integrated, nationwide, digital-health information platform. Databases include all severe acute respiratory syndrome coronavirus 2 (SARS-CoV-2)-related data with no missing information since pandemic onset, such as all polymerase chain reaction (PCR) tests, and from January 5, 2022 onward, all rapid antigen tests conducted at healthcare facilities. SARS-CoV-2 testing in the healthcare system in Qatar is done at a mass scale, and mostly for routine reasons, where about 5% of the population are tested every week (*9, 27*). About 75% of those diagnosed are diagnosed not because of appearance of symptoms, but because of routine testing (*9, 27*). Every PCR test and an increasing proportion of the facility-based rapid antigen tests conducted in Qatar, regardless of location or setting, are classified on the basis of symptoms and the reason for testing (clinical symptoms, contact tracing, surveys or random testing campaigns, individual requests, routine healthcare testing, pre-travel, at port of entry, or other). All facility-based testing done during follow-up in the present study was factored in the analyses of this study.

Rapid antigen test kits are available for purchase in pharmacies in Qatar, but outcome of home-based testing is not reported nor documented in the national databases. Since SARS-CoV-2-test outcomes are linked to specific public health measures, restrictions, and privileges, testing policy and guidelines stress facility-based testing as the core testing mechanism in the population. While facility-based testing is provided free of charge or at low subsidized costs, depending on the reason for testing, home-based rapid antigen testing is de-emphasized and not supported as part of national policy. There is no reason to believe that home-based testing could have differentially affected the followed matched cohorts to affect our results.

The infection detection rate is defined as the cumulative number of documented infections, that is diagnosed and laboratory-confirmed infections, over the cumulative number of documented and undocumented infections. Serological surveys and other analyses suggest that a substantial proportion of infections in Qatar and elsewhere are undocumented (*33-36, 40-42*). With absence of recent serological surveys in Qatar, it is difficult to estimate the current or recent infection detection rate, but mathematical modeling analyses and their recent updates suggest that at present no less than 50% of infections are never documented (*33, 43*).

Differences in testing rate during follow-up may introduce differential ascertainment of infection across the cohorts if routine testing varied by cohort. There was evidence for differences in the testing rate across the cohorts. These differences could result in different rates of undocumented infection before and during follow-up. To address these differences, analyses were conducted by further adjusting the hazard ratios in the Cox regressions for the differences in testing rate (please note below).

Qatar has unusually young, diverse demographics, in that only 9% of its residents are ≥50 years of age, and 89% are expatriates from over 150 countries (*32, 37*). Qatar launched its COVID-19 vaccination program in December of 2020 using the BNT162b2 and mRNA-1273 vaccines (*17*). Detailed descriptions of Qatar’s population and of the national databases have been reported previously (*9, 23, 27, 32, 44*).

### Study design and cohorts

Matched, retrospective, observational cohort studies were conducted to investigate epidemiological evidence for immune imprinting in individuals who had a documented primary omicron infection, but different prior vaccination histories. A documented primary omicron infection was defined as the first record of a SARS-CoV-2-positive PCR or rapid antigen test after onset of the omicron wave in Qatar on December 19, 2021 (*6*) in an individual that had no record of a prior pre-omicron infection.

In the first study, we compared incidence of reinfection in the national cohort of individuals who had a primary omicron infection after primary-series (two-dose) vaccination (designated as the two-dose cohort) to that in the national cohort of individuals who had a primary omicron infection, but no vaccination history (designated as the unvaccinated cohort).

In the second study, we compared incidence of reinfection in the national cohort of individuals who had a primary omicron infection after booster (third dose) vaccination (designated as the three-dose cohort) to that in the two-dose cohort. In a third study, to confirm and complement results of the first two studies, we compared incidence of reinfection in the three-dose cohort to that in the unvaccinated cohort. The majority of primary omicron infections in these three studies involved the BA.2 subvariant (*9-11*).

SARS-CoV-2 reinfection was defined as a documented infection ≥90 days after an earlier infection, to avoid misclassifying prolonged positivity as reinfection (*6-8*). Children vaccinated with the pediatric dose of BNT162b2 and adults who received different vaccines were excluded. Classification of infection severity followed World Health Organization (WHO) guidelines for COVID-19 case severity (acute-care hospitalizations) (*45*), criticality (intensive-care-unit hospitalizations) (*45*), and fatality (*46*).

### Cohort matching and follow-up

Cohorts were matched exactly one-to-one by sex, 10-year age group, nationality, and number of chronic coexisting conditions (none, one, two, three or more comorbid conditions) to balance observed confounders between exposure groups that are related to infection risk in Qatar (*32-36*). Individuals who were first diagnosed with SARS-CoV-2 in a specific week in one cohort were matched to individuals who were first diagnosed with SARS-CoV-2 in that same calendar week in the comparator cohort, to ensure that matched pairs were exposed to the same omicron subvariants and had presence in Qatar at the same time. Cohorts were also matched exactly by testing method (PCR versus rapid antigen testing) and by reason for testing for the primary omicron infection to control for potential differences in testing modalities between cohorts.

Matching was performed iteratively such that individuals in the comparator cohort were alive, had not been reinfected, and had maintained the same vaccination status at the start of follow-up. Each matched pair was followed from 90 days after the primary omicron infection of the individual in the two-dose cohort for the study comparing incidence of reinfection in that cohort with the unvaccinated cohort. Follow-up was from 90 days after the primary omicron infection of the individual in the three-dose cohort for studies comparing incidence of reinfection in that cohort to that in each of the two-dose and unvaccinated cohorts.

For exchangeability (*23, 47*), both members of each matched pair were censored as soon as one of them received a new vaccine dose (change in vaccination status; that is at earliest occurrence of an unvaccinated individual in the matched pair receiving the first dose, or the individual with two-dose vaccination receiving a third dose, or the individual with three-dose vaccination receiving a fourth dose). Accordingly, individuals were followed up until the first of any of the following events: a documented SARS-CoV-2 reinfection (defined as the first PCR-positive or rapid-antigen-positive test after the start of follow-up, regardless of symptoms), a change in vaccination status (with matched-pair censoring), or death, or end of study censoring (September 15, 2022).

### Comorbidity classification

Comorbidities were ascertained and classified based on the ICD-10 codes for chronic conditions as recorded in the electronic health record encounters of each individual in the Cerner-system national database that includes all citizens and residents registered in the national and universal public healthcare system. The public healthcare system provides healthcare to the entire resident population of Qatar free of charge or at heavily subsidized costs, including prescription drugs. With the mass expansion of this sector in recent years, facilities have been built to cater to specific needs of subpopulations. For example, tens of facilities have been built, including clinics and hospitals, in localities with high density of craft and manual workers (*35*).

All encounters for each individual were analyzed to determine the comorbidity classification for that individual, including also all laboratory data, as part of a recent national analysis to assess healthcare needs and resource allocation. The Cerner-system national database includes encounters starting from 2013, after this system was launched in Qatar. As long as each individual had at least one encounter with a specific comorbidity diagnosis based on clinical and laboratory data since 2013, this person was classified with this comorbidity.

Individuals who have comorbidities but never sought care in the public healthcare system, or seek care exclusively in private healthcare facilities, were classified as individuals with no comorbidity due to absence of recorded encounters for them.

It is unlikely that the approach for dealing with coexisting conditions (or implicitly current medications) is of consequence on the results. The population of Qatar is young, of working age, and healthy and the number of persons with severe or multiple chronic conditions is small (*32, 37*). The national list of vaccine prioritization included only 19,800 individuals of all age groups with serious co-morbid conditions to be prioritized in the first phase of vaccine roll-out (Qatar has a total population of about 3 million people) (*27*).

### Laboratory methods and variant ascertainment

#### Real-time reverse-transcription polymerase chain reaction testing

Nasopharyngeal and/or oropharyngeal swabs were collected for PCR testing and placed in Universal Transport Medium (UTM). Aliquots of UTM were: 1) extracted on KingFisher Flex (Thermo Fisher Scientific, USA), MGISP-960 (MGI, China), or ExiPrep 96 Lite (Bioneer, South Korea) followed by testing with real-time reverse-transcription PCR (RT-qPCR) using TaqPath COVID-19 Combo Kits (Thermo Fisher Scientific, USA) on an ABI 7500 FAST (Thermo Fisher Scientific, USA); 2) tested directly on the Cepheid GeneXpert system using the Xpert Xpress SARS-CoV-2 (Cepheid, USA); or 3) loaded directly into a Roche cobas 6800 system and assayed with the cobas SARS-CoV-2 Test (Roche, Switzerland). The first assay targets the viral S, N, and ORF1ab gene regions. The second targets the viral N and E-gene regions, and the third targets the ORF1ab and E-gene regions.

All PCR testing was conducted at the Hamad Medical Corporation Central Laboratory or Sidra Medicine Laboratory, following standardized protocols.

#### Rapid antigen testing

SARS-CoV-2 antigen tests were performed on nasopharyngeal swabs using one of the following lateral flow antigen tests: Panbio COVID-19 Ag Rapid Test Device (Abbott, USA); SARS-CoV-2 Rapid Antigen Test (Roche, Switzerland); Standard Q COVID-19 Antigen Test (SD Biosensor, Korea); or CareStart COVID-19 Antigen Test (Access Bio, USA). All antigen tests were performed point-of-care according to each manufacturer’s instructions at public or private hospitals and clinics throughout Qatar with prior authorization and training by the Ministry of Public Health (MOPH). Antigen test results were electronically reported to the MOPH in real time using the Antigen Test Management System which is integrated with the national COVID-19 database.

#### Classification of infections by variant type

Surveillance for SARS-CoV-2 variants in Qatar is based on viral genome sequencing and multiplex real-time reverse-transcription PCR (RT-qPCR) variant screening (*48*) of random positive clinical samples (*27, 39, 49-52*), complemented by deep sequencing of wastewater samples (*50, 53, 54*). Further details on the viral genome sequencing and multiplex RT-qPCR variant screening throughout the SARS-CoV-2 waves in Qatar can be found in previous publications (*6, 9, 10, 12, 23, 27, 39, 49-52, 55-57*).

### COVID-19 severity, criticality, and fatality classification

Classification of COVID-19 case severity (acute-care hospitalizations) (*45*), criticality (intensive-care-unit hospitalizations) (*45*), and fatality (*46*) followed WHO guidelines. Assessments were made by trained medical personnel independent of study investigators and using individual chart reviews, as part of a national protocol applied to every hospitalized COVID-19 patient. Each hospitalized COVID-19 patient underwent an infection severity assessment every three days until discharge or death. We classified individuals who progressed to severe, critical, or fatal COVID-19 between the time of the documented infection and the end of the study based on their worst outcome, starting with death (*46*), followed by critical disease (*45*), and then severe disease (*45*).

Severe COVID-19 disease was defined per WHO classification as a SARS-CoV-2 infected person with “oxygen saturation of <90% on room air, and/or respiratory rate of >30 breaths/minute in adults and children >5 years old (or ≥60 breaths/minute in children <2 months old or ≥50 breaths/minute in children 2-11 months old or ≥40 breaths/minute in children 1–5 years old), and/or signs of severe respiratory distress (accessory muscle use and inability to complete full sentences, and, in children, very severe chest wall indrawing, grunting, central cyanosis, or presence of any other general danger signs)” (*45*). Detailed WHO criteria for classifying Severe acute respiratory syndrome coronavirus 2 (SARS-CoV-2) infection severity can be found in the WHO technical report (*45*).

Critical COVID-19 disease was defined per WHO classification as a SARS-CoV-2 infected person with “acute respiratory distress syndrome, sepsis, septic shock, or other conditions that would normally require the provision of life sustaining therapies such as mechanical ventilation (invasive or non-invasive) or vasopressor therapy” (*45*). Detailed WHO criteria for classifying SARS-CoV-2 infection criticality can be found in the WHO technical report (*45*).

COVID-19 death was defined per WHO classification as “a death resulting from a clinically compatible illness, in a probable or confirmed COVID-19 case, unless there is a clear alternative cause of death that cannot be related to COVID-19 disease (e.g. trauma).

There should be no period of complete recovery from COVID-19 between illness and death. A death due to COVID-19 may not be attributed to another disease (e.g. cancer) and should be counted independently of preexisting conditions that are suspected of triggering a severe course of COVID-19”. Detailed WHO criteria for classifying COVID-19 death can be found in the WHO technical report (*46*).

### Oversight

The institutional review boards at Hamad Medical Corporation and Weill Cornell Medicine–Qatar approved this retrospective study with a waiver of informed consent. The study was reported according to the Strengthening the Reporting of Observational Studies in Epidemiology (STROBE) guidelines (Table S3). The authors vouch for the accuracy and completeness of the data and for the fidelity of the study to the protocol. Data used in this study are the property of the Ministry of Public Health of Qatar and were provided to the researchers through a restricted-access agreement for preservation of confidentiality of patient data. The funders had no role in the study design; the collection, analysis, or interpretation of the data; or the writing of the manuscript.

### Statistical analysis

Eligible and matched cohorts were drawn from independent samples and described using frequency distributions and measures of central tendency and were compared using standardized mean differences (SMDs). An SMD of ≤0.1 indicated adequate matching (*58*). Cumulative incidence of reinfection (defined as proportion of individuals at risk, whose primary endpoint during follow-up was a reinfection) was estimated using the Kaplan-Meier estimator method (*59*). Incidence rate of reinfection in each cohort, defined as number of identified reinfections divided by number of person-weeks contributed by all individuals in the cohort, was estimated, with the corresponding 95% confidence interval (CI) using a Poisson log-likelihood regression model with the Stata 17.0 *stptime* command.

Hazard ratios, comparing incidence of reinfection in the cohorts and corresponding 95% CIs, were calculated using Cox regression, adjusted for the matching factors with the Stata 17.0 *stcox* command. The overall hazard ratio and the month-by-month hazard ratios in the Cox regression were additionally adjusted for differences in testing rate (low testers, intermediate testers, and high testers defined as persons having ≤2, 3-6, and ≥7 tests per person-year during follow-up, respectively). This additional adjustment was conducted because most SARS-CoV-2 testing in Qatar is done for routine reasons and not because of symptoms (*9, 27*). About 75% of those diagnosed with the infection are diagnosed not because of appearance of symptoms, but because of routine testing (*9, 27*). Testing guidelines also differed by vaccination status (such as for travel-related testing) (*27*). Any differences in testing rate can potentially introduce differential ascertainment of infection across the cohorts if routine testing varied by cohort.

Sensitivity analyses were also conducted for the central analysis comparing incidence of reinfection in the three-dose cohort to the two-dose cohort. In the first analysis, the cohorts were matched by the Charlson comorbidity index instead of the number of coexisting conditions. In the second analysis, the cohorts were matched additionally by primary-series vaccine type (two doses of BNT162b2 or two doses of mRNA-1273). Subgroup analyses were also conducted for the latter sensitivity analysis where the hazard ratios were calculated separately for each of BNT162b2- and mRNA-1273-vaccinated individuals.

Schoenfeld residuals and log-log plots for survival curves were used to test the proportional-hazards assumption. CIs were not adjusted for multiplicity; thus, they should not be used to infer definitive differences between groups. Interactions were not considered. Statistical analyses were conducted using Stata/SE version 17.0 (Stata Corporation, College Station, TX, USA).

## Data Availability

All relevant data needed to evaluate the conclusions in the paper are present in the paper and/or the Supplementary Materials. The raw dataset of this study is a property of the Qatar Ministry of Public Health that was provided to the researchers through a restricted-access agreement that prevents sharing the dataset with a third party or publicly. The data are available under restricted access for preservation of confidentiality of patient data. Access can be obtained through a direct application for data access to Her Excellency the Minister of Public. The raw data are protected and are not available due to data privacy laws. Data were available to authors through.csv files where information has been downloaded from the CERNER database system (no links/accession codes were available o authors).

## Acknowledgments

We acknowledge the many dedicated individuals at Hamad Medical Corporation, the Ministry of Public Health, the Primary Health Care Corporation, Qatar Biobank, Sidra Medicine, and Weill Cornell Medicine-Qatar for their diligent efforts and contributions to make this study possible. The authors are grateful for institutional salary support from the Biomedical Research Program and the Biostatistics, Epidemiology, and Biomathematics Research Core, both at Weill Cornell Medicine-Qatar, as well as for institutional salary support provided by the Ministry of Public Health, Hamad Medical Corporation, and Sidra Medicine. The authors are also grateful for the Qatar Genome Programme and Qatar University Biomedical Research Center for institutional support for the reagents needed for the viral genome sequencing. HHA acknowledges the support of Qatar University collaborative grant QUCG-CAS-23/24-114. The funders of the study had no role in study design, data collection, data analysis, data interpretation, or writing of the article. Statements made herein are solely the responsibility of the authors.

## Funding

Biomedical Research Program and the Biostatistics, Epidemiology, and Biomathematics Research Core, both at Weill Cornell Medicine-Qatar Ministry of Public Health, Hamad Medical Corporation, and Sidra Medicine Qatar Genome Programme and Qatar University Biomedical Research Center Qatar University collaborative grant QUCG-CAS-23/24-114.

## Author contributions

Conceptualization: LJA, HC

Data Curation: HC, ZAK, EAK, AJ, AHK, ANL, RMS, MAK, AAB, HEAR, MHAT, AAK, RB, and LJA

Methodology: LJA, HC

Investigation: LJA, HC, HHA, PT, MRH, PVC, HY, AAT, HAK

Visualization: LJA

Funding acquisition: LJA

Project administration: LJA

Supervision: LJA

Writing – original draft: LJA, HC

Writing – review & editing: LJA, HC, HHA, PT, PVC, HY, AAT, HAK, MRH, ZAK, EAK, AJ, AHK, ANL, RMS, HAR, GKN, MAK, AAB, HEAR, MHAT, AAK, RB

## Competing interests

Dr. Butt has received institutional grant funding from Gilead Sciences unrelated to the work presented in this paper. Otherwise, we declare no competing interests.

## Supplementary Materials for

**Fig. S1.**
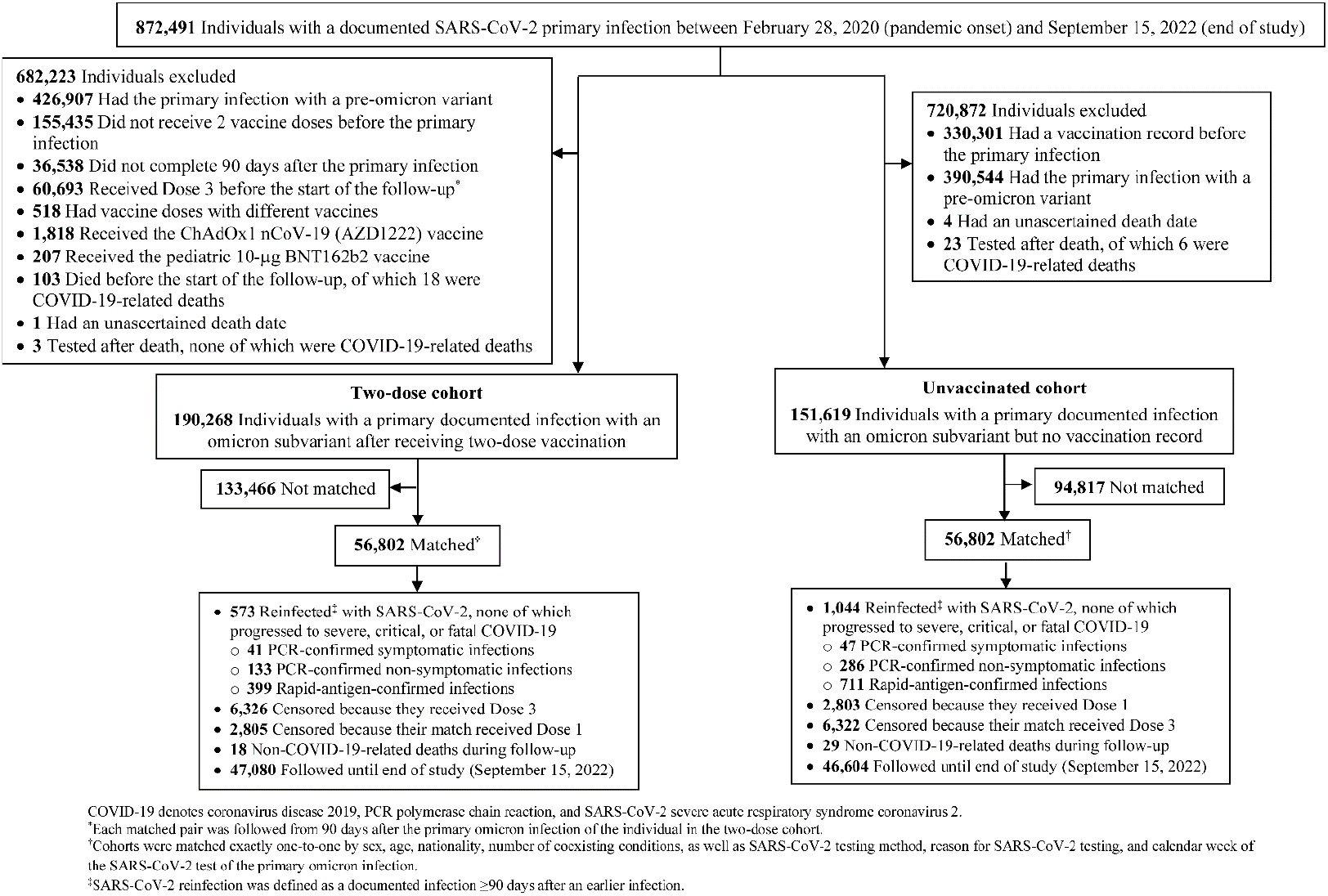
Flowchart describing the population selection process for investigating immune protection against reinfection among those who had a primary infection with an omicron subvariant after two-dose vaccination compared to protection among those who had a primary infection with an omicron subvariant but were unvaccinated.

**Fig. S2.**
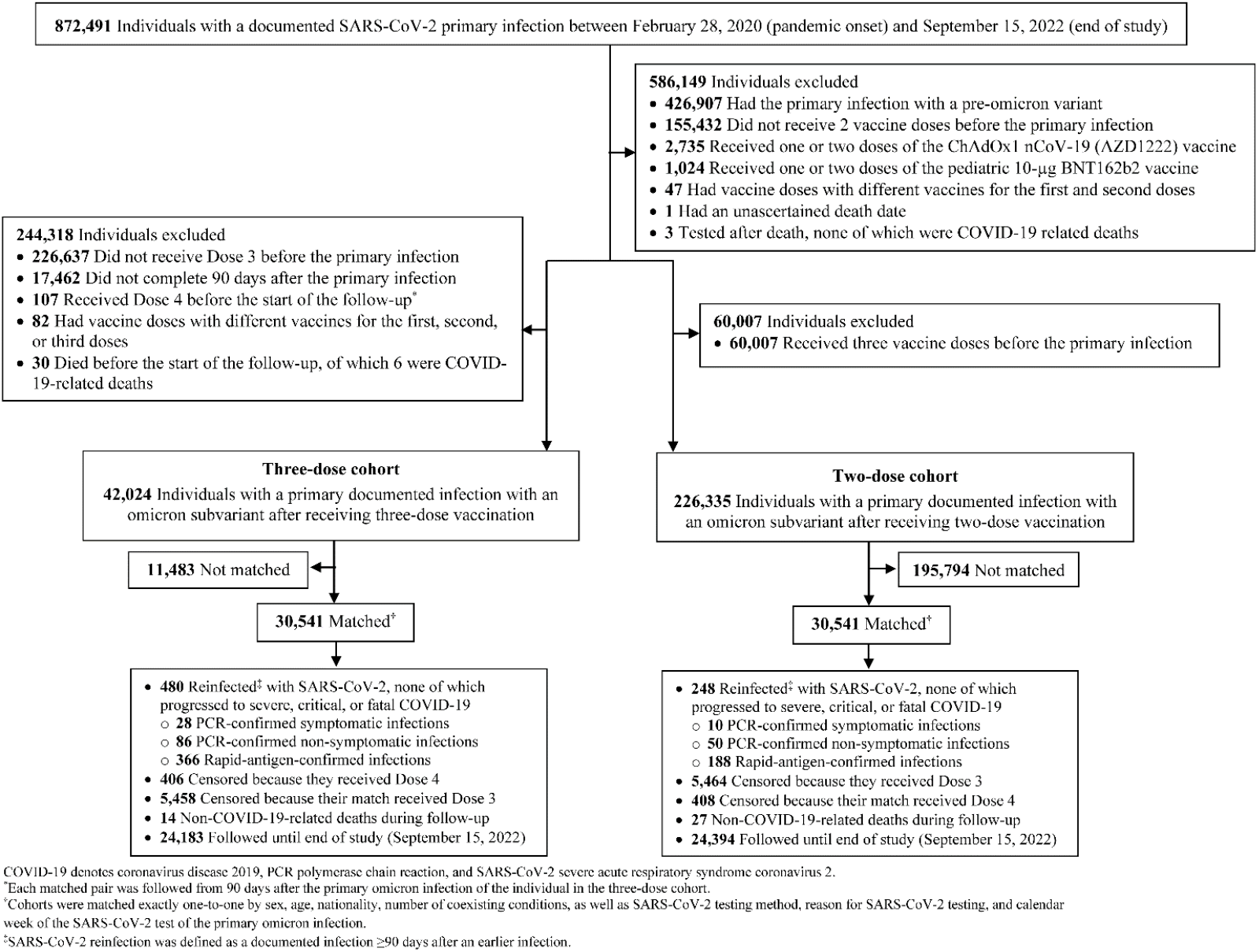
Flowchart describing the population selection process for investigating immune protection against reinfection among those who had a primary infection with an omicron subvariant after three-dose vaccination compared to protection among those who had a primary infection with an omicron subvariant after two-dose vaccination.

**Table S1.** Sensitivity analyses. Hazard ratios for incidence of SARS-CoV-2 reinfection in the study investigating immune protection among those who had a primary infection with an omicron subvariant after three-dose vaccination compared to two-dose vaccination.

| Epidemiological measure | Cohorts |  |
| --- | --- | --- |
| A) Matching by Charlson comorbidity index score<sup>a</sup> | Three-dose cohort | Two-dose cohort |
| Sample size | 29,508 | 29,508 |
| Incident reinfections (n) | 478 | 236 |
| Total follow-up time (person-weeks) | 566,587 | 568,088 |
| Incidence rate of reinfection (per 10,000 person-weeks; 95% CI) | 8.4 (7.7 to 9.2) | 4.2 (3.7 to 4.7) |
| Unadjusted hazard ratio for SARS-CoV-2 reinfection (95% CI) | 2.04 (1.74 to 2.38) |  |
| Adjusted hazard ratio for SARS-CoV-2 reinfection (95% CI)<sup>b</sup> | 1.99 (1.67 to 2.39) |  |
| Hazard ratio for SARS-CoV-2 reinfection additionally adjusted for differences in testing frequency (95% CI)<sup>b</sup> | 1.39 (1.16 to 1.67) |  |
| B) Matching by primary-series vaccine type<sup>c</sup> | Three-dose cohort | Two-dose cohort |
| Sample size | 28,357 | 28,357 |
| Incident reinfections (n) | 453 | 243 |
| Total follow-up time (person-weeks) | 546,890 | 548,008 |
| Incidence rate of reinfection (per 10,000 person-weeks; 95% CI) | 8.3 (7.6 to 9.1) | 4.4 (3.9 to 5.0) |
| Unadjusted hazard ratio for SARS-CoV-2 reinfection (95% CI) | 1.87 (1.60 to 2.19) |  |
| Adjusted hazard ratio for SARS-CoV-2 reinfection (95% CI)<sup>d</sup> | 1.94 (1.62 to 2.33) |  |
| Hazard ratio for SARS-CoV-2 reinfection additionally adjusted for differences in testing frequency (95% CI)<sup>d</sup> | 1.43 (1.19 to 1.71) |  |
| Cohorts who received BNT162b2 primary series | Three-dose cohort | Two-dose cohort |
| Sample size | 23,533 | 23,533 |
| Incident reinfections (n) | 405 | 223 |
| Total follow-up time (person-weeks) | 456,085 | 457,065 |
| Incidence rate of reinfection (per 10,000 person-weeks; 95% CI) | 8.9 (8.1 to 9.8) | 4.9 (4.3 to 5.6) |
| Unadjusted hazard ratio for SARS-CoV-2 reinfection (95% CI) | 1.82 (1.55 to 2.15) |  |
| Adjusted hazard ratio for SARS-CoV-2 reinfection (95% CI) | 1.89 (1.56 to 2.29) |  |
| Hazard ratio for SARS-CoV-2 reinfection additionally adjusted for differences in testing frequency (95% CI) | 1.39 (1.15 to 1.68) |  |
| Cohorts who received mRNA-1273 primary series |  |  |
| Sample size | 4,804 | 4,804 |
| Incident reinfections (n) | 48 | 20 |
| Total follow-up time (person-weeks) | 90,805 | 90,943 |
| Incidence rate of reinfection (per 10,000 person-weeks; 95% CI) | 5.3 (4.0 to 7.0) | 2.2 (1.4 to 3.4) |
| Unadjusted hazard ratio for SARS-CoV-2 reinfection (95% CI) | 2.41 (1.43 to 4.06) |  |
| Adjusted hazard ratio for SARS-CoV-2 reinfection (95% CI) | 2.45 (1.37 to 4.39) |  |
| Hazard ratio for SARS-CoV-2 reinfection additionally adjusted for differences in testing frequency (95% CI) | 1.83 (1.03 to 3.28) |  |

CI denotes confidence interval and SARS-CoV-2 severe acute respiratory syndrome coronavirus 2.
<sup>a</sup>Cohorts were matched exactly one-to-one by sex, age, nationality, Charlson comorbidity index, as well as SARS-CoV-2 testing method, reason for SARS-CoV-2 testing, and calendar week of the SARS-CoV-2 test of the primary omicron infection.
<sup>b</sup>Cox regression analysis adjusted for sex, 10-year age groups, 10 nationality groups, Charlson comorbidity index, as well as SARS-CoV-2 testing method, reason for SARS-CoV-2 testing, and calendar week of the SARS-CoV-2 test of the primary omicron infection.
<sup>c</sup>Cohorts were matched exactly one-to-one by sex, age, nationality, number of coexisting conditions, primary-series vaccine type (two doses of BNT162b2 or two doses of mRNA-1273), as well as SARS-CoV-2 testing method, reason for SARS-CoV-2 testing, and calendar week of the SARS-CoV-2 test of the primary omicron infection.
<sup>d</sup>Cox regression analysis adjusted for sex, 10-year age groups, 10 nationality groups, number of coexisting conditions, primary-series vaccine type, as well as SARS-CoV-2 testing method, reason for SARS-CoV-2 testing, and calendar week of the SARS-CoV-2 test of the primary omicron infection.

**Fig. S3.**
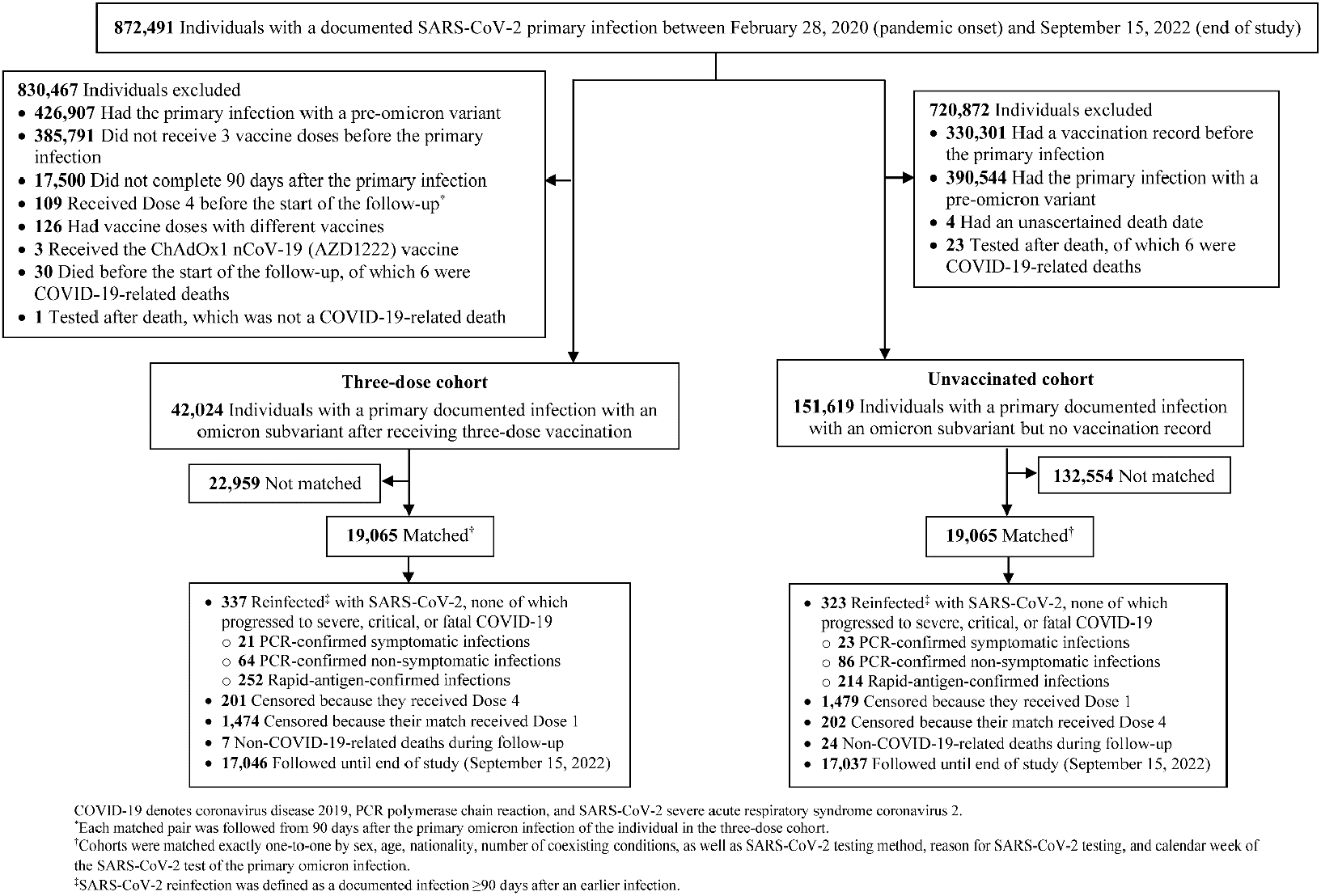
Flowchart describing the population selection process for investigating immune protection against reinfection among those who had a primary infection with an omicron subvariant after three-dose vaccination compared to protection among those who had a primary infection with an omicron subvariant but were unvaccinated.

**Table S2.**
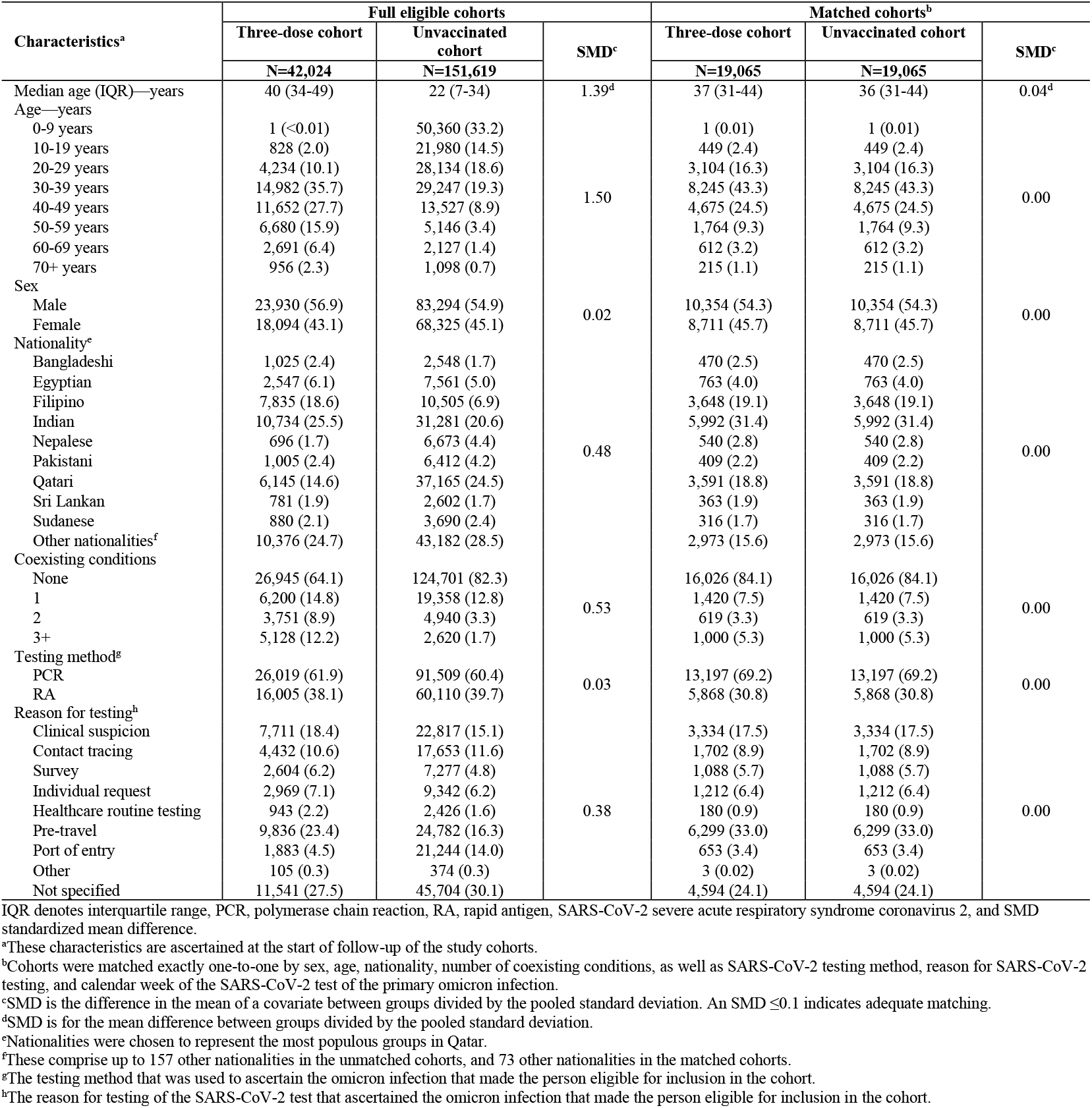
Baseline characteristics of eligible and matched cohorts in the study investigating immune protection against reinfection among those who had a primary infection with an omicron subvariant after three-dose vaccination compared to those who had a primary infection with an omicron subvariant but were unvaccinated.

| Characteristics<sup>a</sup> | Full eligible cohorts |  |  | Matched cohorts<sup>b</sup> |  |  |
| --- | --- | --- | --- | --- | --- | --- |
|  | Three-dose cohort | Unvaccinated cohort | SMD<sup>c</sup> | Three-dose cohort | Unvaccinated cohort | SMD<sup>c</sup> |
|  | N=42,024 | N=151,619 |  | N=19,065 | N=19,065 |  |
| Median age (IQR)—years | 40 (34-49) | 22 (7-34) | 1.39<sup>d</sup> | 37 (31-44) | 36 (31-44) | 0.04<sup>d</sup> |
| Age—years |  |  |  |  |  |  |
| 0-9 years | 1 (<0.01) | 50,360 (33.2) |  | 1 (0.01) | 1 (0.01) |  |
| 10-19 years | 828 (2.0) | 21,980 (14.5) |  | 449 (2.4) | 449 (2.4) |  |
| 20-29 years | 4,234 (10.1) | 28,134 (18.6) |  | 3,104 (16.3) | 3,104 (16.3) |  |
| 30-39 years | 14,982 (35.7) | 29,247 (19.3) | 1.50 | 8,245 (43.3) | 8,245 (43.3) | 0.00 |
| 40-49 years | 11,652 (27.7) | 13,527 (8.9) |  | 4,675 (24.5) | 4,675 (24.5) |  |
| 50-59 years | 6,680 (15.9) | 5,146 (3.4) |  | 1,764 (9.3) | 1,764 (9.3) |  |
| 60-69 years | 2,691 (6.4) | 2,127 (1.4) |  | 612 (3.2) | 612 (3.2) |  |
| 70+ years | 956 (2.3) | 1,098 (0.7) |  | 215 (1.1) | 215 (1.1) |  |
| Sex |  |  |  |  |  |  |
| Male | 23,930 (56.9) | 83,294 (54.9) | 0.02 | 10,354 (54.3) | 10,354 (54.3) | 0.00 |
| Female | 18,094 (43.1) | 68,325 (45.1) |  | 8,711 (45.7) | 8,711 (45.7) |  |
| Nationality<sup>e</sup> |  |  |  |  |  |  |
| Bangladeshi | 1,025 (2.4) | 2,548 (1.7) |  | 470 (2.5) | 470 (2.5) |  |
| Egyptian | 2,547 (6.1) | 7,561 (5.0) |  | 763 (4.0) | 763 (4.0) |  |
| Filipino | 7,835 (18.6) | 10,505 (6.9) |  | 3,648 (19.1) | 3,648 (19.1) |  |
| Indian | 10,734 (25.5) | 31,281 (20.6) |  | 5,992 (31.4) | 5,992 (31.4) |  |
| Nepalese | 696 (1.7) | 6,673 (4.4) | 0.48 | 540 (2.8) | 540 (2.8) | 0.00 |
| Pakistani | 1,005 (2.4) | 6,412 (4.2) |  | 409 (2.2) | 409 (2.2) |  |
| Qatari | 6,145 (14.6) | 37,165 (24.5) |  | 3,591 (18.8) | 3,591 (18.8) |  |
| Sri Lankan | 781 (1.9) | 2,602 (1.7) |  | 363 (1.9) | 363 (1.9) |  |
| Sudanese | 880 (2.1) | 3,690 (2.4) |  | 316 (1.7) | 316 (1.7) |  |
| Other nationalities<sup>f</sup> | 10,376 (24.7) | 43,182 (28.5) |  | 2,973 (15.6) | 2,973 (15.6) |  |
| Coexisting conditions |  |  |  |  |  |  |
| None | 26,945 (64.1) | 124,701 (82.3) |  | 16,026 (84.1) | 16,026 (84.1) |  |
| 1 | 6,200 (14.8) | 19,358 (12.8) | 0.53 | 1,420 (7.5) | 1,420 (7.5) | 0.00 |
| 2 | 3,751 (8.9) | 4,940 (3.3) |  | 619 (3.3) | 619 (3.3) |  |
| 3+ | 5,128 (12.2) | 2,620 (1.7) |  | 1,000 (5.3) | 1,000 (5.3) |  |
| Testing method<sup>g</sup> |  |  |  |  |  |  |
| PCR | 26,019 (61.9) | 91,509 (60.4) | 0.03 | 13,197 (69.2) | 13,197 (69.2) | 0.00 |
| RA | 16,005 (38.1) | 60,110 (39.7) |  | 5,868 (30.8) | 5,868 (30.8) |  |
| Reason for testing<sup>h</sup> |  |  |  |  |  |  |
| Clinical suspicion | 7,711 (18.4) | 22,817 (15.1) |  | 3,334 (17.5) | 3,334 (17.5) |  |
| Contact tracing | 4,432 (10.6) | 17,653 (11.6) |  | 1,702 (8.9) | 1,702 (8.9) |  |
| Survey | 2,604 (6.2) | 7,277 (4.8) |  | 1,088 (5.7) | 1,088 (5.7) |  |
| Individual request | 2,969 (7.1) | 9,342 (6.2) |  | 1,212 (6.4) | 1,212 (6.4) |  |
| Healthcare routine testing | 943 (2.2) | 2,426 (1.6) | 0.38 | 180 (0.9) | 180 (0.9) | 0.00 |
| Pre-travel | 9,836 (23.4) | 24,782 (16.3) |  | 6,299 (33.0) | 6,299 (33.0) |  |
| Port of entry | 1,883 (4.5) | 21,244 (14.0) |  | 653 (3.4) | 653 (3.4) |  |
| Other | 105 (0.3) | 374 (0.3) |  | 3 (0.02) | 3 (0.02) |  |
| Not specified | 11,541 (27.5) | 45,704 (30.1) |  | 4,594 (24.1) | 4,594 (24.1) |  |

IQR denotes interquartile range, PCR, polymerase chain reaction, RA, rapid antigen, SARS-CoV-2 severe acute respiratory syndrome coronavirus 2, and SMD standardized mean difference.
<sup>a</sup>These characteristics are ascertained at the start of follow-up of the study cohorts.
<sup>b</sup>Cohorts were matched exactly one-to-one by sex, age, nationality, number of coexisting conditions, as well as SARS-CoV-2 testing method, reason for SARS-CoV-2 testing, and calendar week of the SARS-CoV-2 test of the primary omicron infection.
<sup>c</sup>SMD is the difference in the mean of a covariate between groups divided by the pooled standard deviation. An SMD $\leq 0.1$ indicates adequate matching.
<sup>d</sup>SMD is for the mean difference between groups divided by the pooled standard deviation.
<sup>e</sup>Nationalities were chosen to represent the most populous groups in Qatar.
<sup>f</sup>These comprise up to 157 other nationalities in the unmatched cohorts, and 73 other nationalities in the matched cohorts.
<sup>g</sup>The testing method that was used to ascertain the omicron infection that made the person eligible for inclusion in the cohort.
<sup>h</sup>The reason for testing of the SARS-CoV-2 test that ascertained the omicron infection that made the person eligible for inclusion in the cohort.

**Fig. S4.**
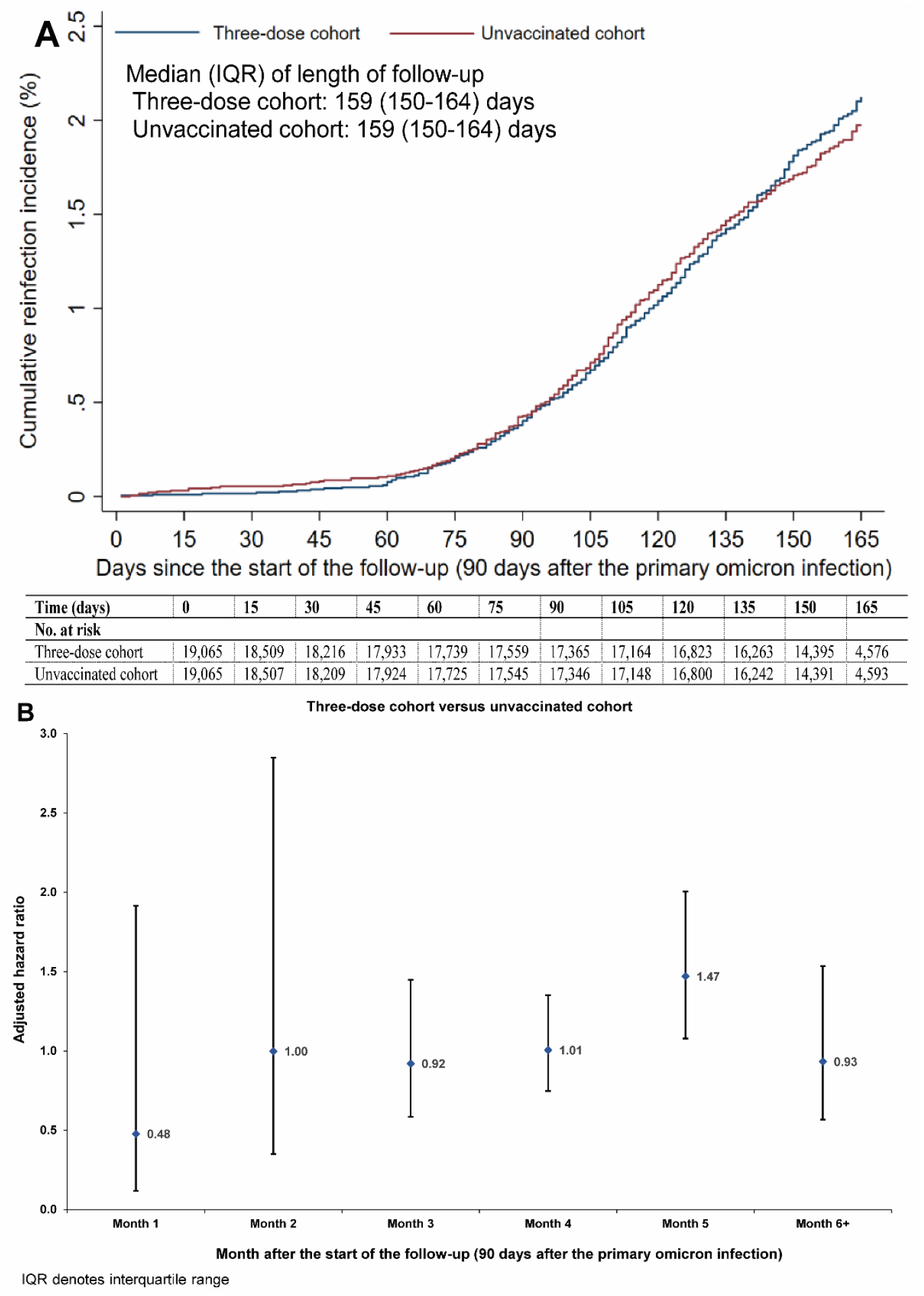
A) Cumulative incidence of and B) adjusted hazard ratio by month of follow-up for SARS-CoV-2 reinfection among those who had a primary infection with an omicron subvariant after three-dose vaccination compared to those who had a primary infection with an omicron subvariant but were unvaccinated. Error bars indicate confidence intervals.

**Fig. S5.**
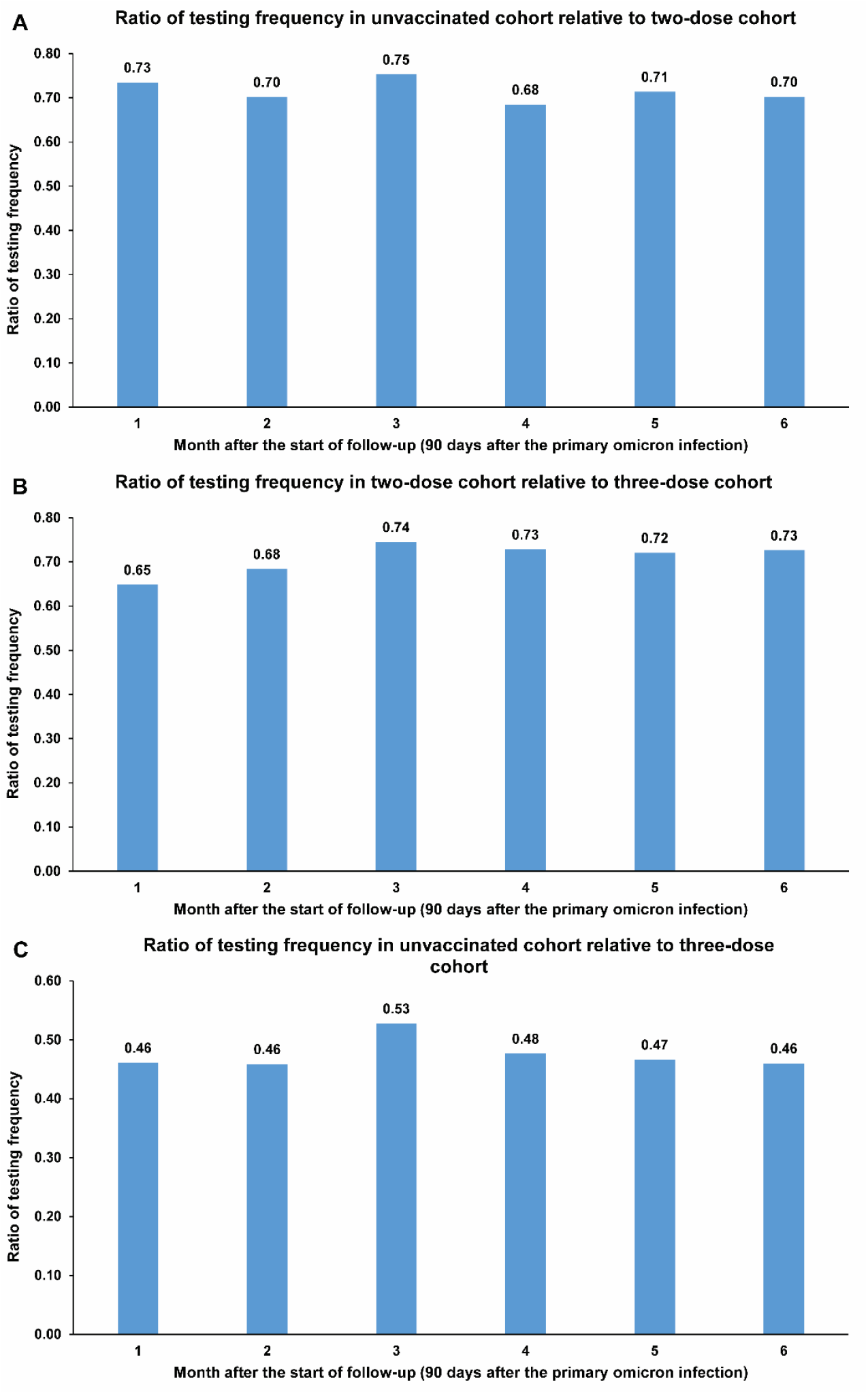
Ratio of testing frequency in the matched cohorts of studies investigating immune protection among those who had a primary infection with an omicron subvariant, but different vaccination histories.

**Table S3.** STROBE checklist for cohort studies.

|  | Item No | Recommendation | Main Text page |
| --- | --- | --- | --- |
| Title and abstract | 1 | (a) Indicate the study’s design with a commonly used term in the title or the abstract<br>(b) Provide in the abstract an informative and balanced summary of what was done and what was found | Abstract<br>Abstract |
| Introduction |  |  |  |
| Background/rationale | 2 | Explain the scientific background and rationale for the investigation being reported | Introduction |
| Objectives | 3 | State specific objectives, including any prespecified hypotheses | Introduction |
| Methods |  |  |  |
| Study design | 4 | Present key elements of study design early in the paper | Intoduction (paragraph 4) & Materials & Methods (‘Study design and cohorts’ & ‘Cohort matching and follow-up’) |
| Setting | 5 | Describe the setting, locations, and relevant dates, including periods of recruitment, exposure, follow-up, and data collection | Materials & Methods (‘Study population and data sources’, ‘Study design and cohorts’ & ‘Cohort matching and follow-up’, & Figs. S1-S3 |
| Participants | 6 | (a) Give the eligibility criteria, and the sources and methods of selection of participants. Describe methods of follow-up<br>(b) For matched studies, give matching criteria and number of exposed and unexposed | Materials & Methods (‘Study design and cohorts’ & ‘Cohort matching and follow-up’, & Figs. S1-S3 |
| Variables | 7 | Clearly define all outcomes, exposures, predictors, potential confounders, and effect modifiers. Give diagnostic criteria, if applicable | Materials & Methods (‘Study design and cohorts’ & ‘Cohort matching and follow-up’, ‘Comorbidity classification’, ‘Laboratory methods’, ‘COVID-19 severity, criticality, and fatality classification’), Table 1, & Table S2 |
| Data sources/measurement | 8* | For each variable of interest, give sources of data and details of methods of assessment (measurement). Describe comparability of assessment methods if there is more than one group | Materials & Methods (‘Study population and data sources’, ‘Comorbidity classification’, ‘Laboratory methods’, ‘COVID-19 severity, criticality, and fatality classification’, & ‘Statistical analysis’, paragraph 1), Table 1, & Table S2 |
| Bias | 9 | Describe any efforts to address potential sources of bias | Materials & Methods (‘Cohort matching and follow-up’ & ‘Statistical analysis’, paragraph 2) |
| Study size | 10 | Explain how the study size was arrived at | Figs. S1-S3 |
| Quantitative variables | 11 | Explain how quantitative variables were handled in the analyses. If applicable, describe which groupings were chosen and why | Materials & Methods (‘Cohort matching and follow-up’ & ‘Statistical analysis’, paragraph 2), Table 1, & Table S2 |
| Statistical methods | 12 | (a) Describe all statistical methods, including those used to control for confounding | Materials & Methods (‘Statistical analysis’) |
|  |  | (b) Describe any methods used to examine subgroups and interactions | Materials & Methods (‘Statistical analysis’, paragraphs 2-3) |
|  |  | (c) Explain how missing data were addressed | Not applicable, see Materials & Methods (‘Study population and data sources’) |
|  |  | (d) If applicable, explain how loss to follow-up was addressed | Not applicable, see Materials & Methods (‘Study population and data sources’) |
|  |  | (e) Describe any sensitivity analyses | Materials & Methods (‘Statistical analysis’, paragraph 3) |
| Results |  |  |  |
| Participants | 13* | (a) Report numbers of individuals at each stage of study—eg numbers potentially eligible, examined for eligibility, confirmed eligible, included in the study, completing follow-up, and analysed<br>(b) Give reasons for non-participation at each stage<br>(c) Consider use of a flow diagram | Figs. S1-S3 |
| Descriptive data | 14 | (a) Give characteristics of study participants (eg demographic, clinical, social) and information on exposures and potential confounders | Results (‘Two-dose cohort versus unvaccinated cohort’, paragraphs 1 & 2, ‘Three-dose cohort versus two-dose cohort’, paragraphs 1 & 2, & ‘Three-dose cohort versus unvaccinated cohort’, paragraph 1). Table 1, & Table S2 |
|  |  | (b) Indicate number of participants with missing data for each variable of interest | Not applicable, see Materials & Methods ('Study population and data sources') |
|  |  | (c) Summarise follow-up time (eg, average and total amount) | Results ('Two-dose cohort versus unvaccinated cohort', paragraph 2, & 'Three-dose cohort versus two-dose cohort', paragraph 2), Fig. 1, Table 2, & Fig. S4A |
| Outcome data | 15 | Report numbers of outcome events or summary measures over time | Results ('Two-dose cohort versus unvaccinated cohort', paragraphs 3 & 4, 'Three-dose cohort versus two-dose cohort', paragraphs 3 & 4, & 'Three-dose cohort versus unvaccinated cohort', paragraph 2), Fig. 1, Table 2, & Fig. S4 |
| Main results | 16 | (a) Give unadjusted estimates and, if applicable, confounder-adjusted estimates and their precision (eg, 95% confidence interval). Make clear which confounders were adjusted for and why they were included | Results ('Two-dose cohort versus unvaccinated cohort', paragraphs 3 & 4, 'Three-dose cohort versus two-dose cohort', paragraphs 3 & 4, & 'Three-dose cohort versus unvaccinated cohort', paragraph 2), Fig. 1, Table 2, & Fig. S4 |
|  |  | (b) Report category boundaries when continuous variables were categorized | Table 1 & Table S2 |
|  |  | (c) If relevant, consider translating estimates of relative risk into absolute risk for a meaningful time period | Not applicable |
| Other analyses | 17 | Report other analyses done—eg analyses of subgroups and interactions, and sensitivity analyses | Results ('Two-dose cohort versus unvaccinated cohort', paragraph 4, 'Three-dose cohort versus two-dose cohort', paragraph 4, & 'Three-dose cohort versus unvaccinated cohort', paragraph 2-3), Fig. 2, Table S1, Fig. S4B, & Fig. S5 |
| Discussion |  |  |  |
| Key results | 18 | Summarise key results with reference to study objectives | Discussion, paragraphs 1-10 |
| Limitations | 19 | Discuss limitations of the study, taking into account sources of potential bias or imprecision. Discuss both direction and magnitude of any potential bias | Discussion, paragraphs 11-16 |
| Interpretation | 20 | Give a cautious overall interpretation of results considering objectives, limitations, multiplicity of analyses, results from similar studies, and other relevant evidence | Discussion, paragraph 17 |
| Generalisability | 21 | Discuss the generalisability (external validity) of the study results | Discussion, paragraphs 11-16 |
| Other information |  |  |  |
| Funding | 22 | Give the source of funding and the role of the funders for the present study and, if applicable, for the original study on which the present article is based | Funding |

## Notes

### Competing Interest Statement

The authors have declared no competing interest.

### Author Declarations

The institutional review boards at Hamad Medical Corporation and Weill Cornell Medicine in Qatar approved this retrospective study with a waiver of informed consent.

### Summary of Updates

The analyses adjusting the measures for the testing rates among the cohorts have been implemented using an improved method. Discussion has been added of recent developments in relation to this area of research and comparison to newly available data.

